# Ethnic and socioeconomic inequalities in stroke risk factors and primary prevention over three decades: the South London Stroke Register cohort study 1995-2024

**DOI:** 10.64898/2026.01.13.26343999

**Authors:** Eva S Emmett, Camila Pantoja-Ruiz, Evelyn Lim, Amal R Khanolkar, Ajay Bhalla, Charles DA Wolfe, Matthew DL O’Connell, Iain J Marshall

## Abstract

**Objectives:** To investigate long-term trends in ethnic and socioeconomic inequalities in pre-stroke risk factors and treatments.

**Design:** population-based cohort study

**Setting:** geographically defined area of London, UK, 1995-2024

**Participants:** 8,515 adults with first stroke, stratified by self-reported ethnicity (62.4% White [N=5,255], 15.5% Black Caribbean [N=1,303], 13.3% Black African [N=1,122]), occupation (60.7% routine/manual occupations [N=3,964]), education (44.6% lower education [N=1,929]), and 10-year cohorts

**Outcome measures:** pre-stroke vascular risk factor diagnoses and prescribed treatments

**Results:** Mean age at stroke was 59.6 years in Black Africans, 68.1 in Black Caribbeans, and 71.8 in White participants. 12.0% of Black Africans’ strokes occurred without pre-stroke risk factor diagnosis (White participants: 6.3%). Compared to their counterparts, Black participants and those with routine/manual occupations or lower education had higher rates of hypertension (adjusted prevalence ratio: Black Caribbean 1.29 [95%CI:1.24 to 1.34], Black African 1.47 [1.40 to 1.53], manual/routine 1.09 [1.05 to 1.13], lower education 1.06 [1.02 to 1.11]), and diabetes (Black Caribbean 2.23 [2.04 to 2.43], Black African 1.92 [1.73 to 2.13], manual/routine 1.23 [1.13 to 1.34], lower education 1.21 [1.09 to 1.37]), but lower rates of atrial fibrillation (Black Caribbean 0.57 [0.48 to 0.68], Black African 0.66 [0.54 to 0.82], lower education 0.78 [0.68 to 0.91]). Ethnic and socioeconomic inequalities in diabetes and hypertension widened over time but narrowed for atrial fibrillation. 36% of strokes occurred in people with at least one diagnosed, but untreated risk factor throughout the study period. Treatment rates were not associated with occupation or education, but Black participants had higher treatment rates for hypertension and diabetes. Adjusting for occupation or education had negligible impact on associations between ethnicity and risk factor diagnosis or treatment.

**Conclusion:** Prevalence of pre-stroke hypertension and diabetes is higher and rising faster in ethnic minority and lower socioeconomic groups. Together with Black people’s younger stroke age and higher likelihood of having stroke without any prior risk factor diagnosis, these findings indicate that targeted and earlier primary prevention efforts are needed.

**Registration:** This study is registered at ClinicalTrials.gov (Identifier: NCT05298436).

## INTRODUCTION

Stroke remains a major cause of morbidity and mortality worldwide and in the UK; yet ∼90% of strokes are associated with ten modifiable vascular risk factors and therefore potentially preventable.^1^ However, despite the availability of evidence-based guidelines and pay-for-performance policies in many countries, including the UK,^2^ risk factor control remains inadequate, due to shortcomings in detection, prescription, medication adherence, and/or monitoring.^3,4^

Ethnic inequalities in stroke incidence persist, significantly driven by inequalities in risk factor prevalence and control,^3,5–8^ and with strokes occurring at younger ages in ethnic minority groups.^9^ In previous studies, Black people had higher rates of hypertension and lower treatment and control rates compared to their White counterparts, even though suboptimal control confers a three times higher stroke risk.^10,11^ An American study described a steeper rise of diabetes in Black compared to White stroke patients.^12^ Conversely, lower atrial fibrillation (AF) prevalence has been reported in Black people, despite a greater burden of AF risk factors, referred to as the “AF paradox”;^13,14^ and anticoagulation remains widely under-used, especially in ethnic minority groups.^15^

Higher risk factor prevalence and lower treatment and control rates have also been reported in lower socioeconomic groups.^16–19^ Due to the intersectionality between ethnicity and socioeconomic status, these characteristics may compound to increase an individual’s risk factor burden and consequent stroke risk.^20,21^

While ethnic and socioeconomic inequalities in vascular risk factors have been investigated, long-term trends in a population-based stroke cohort and the extent to which socioeconomic inequalities drive ethnic inequalities remain understudied.^21,22^ The South London Stroke Register (SLSR) benefits from a large cohort covering three decades and with significant ethnic and socioeconomic diversity. This allows detailed analyses of long-term trends in ethnic and socioeconomic inequalities and their mutual impact. Compared to US studies, this study’s UK setting with its universal, tax-funded healthcare provision adds a different lens to studying health inequalities. Identifying predominant risk factors and suboptimal treatment patterns in specific groups can inform more targeted approaches of vascular health checks, lifestyle campaigns, and risk factor control. Long-term trends in risk factor diagnoses contribute to evaluating past prevention and detection efforts, estimating future trends, and informing future policies.

Using SLSR data, we investigated inequalities in pre-stroke risk factor diagnoses and treatments between ethnic and socioeconomic groups, their mutual impact, and trends over 30 years.

## METHODS

The SLSR is a population-based cohort study of adults with first stroke since 1995 in a defined area of inner-city London. The study area’s population increased from 310,028 in 2001 (63% White, 15% Black African, 9% Black Caribbean, 13% other ethnic groups, hereafter “Other”) to 398,555 in 2021 (52% White, 15% Black African, 7% Black Caribbean, 26% Other).^23^

The methods of the SLSR have been described previously^24^ and are summarized here. Multiple overlapping notification sources were used to enhance case ascertainment, including hospital admissions, outpatient clinics, radiology reports, A&E records, and general practitioners. Capture-recapture models estimated 88% completeness of case ascertainment.^25^ Stroke diagnosis was verified by senior study clinicians using the World Health Organization definition of stroke ICD-10 until March 2022 and ICD-11 thereafter.

### Exposure

Ethnicity was self-reported, stratified as per UK census^23^ and most common groups into White, Black Caribbean, Black African and Other (other Black groups/Asian/mixed/other).

Socioeconomic status (SES) was estimated using the individual-level indicators occupation and education. Occupation was defined as current or most recent occupation (if currently not working/retired) or spouse’s/father’s occupation for stay-at-home spouses/students. Until 2007, occupations were categorized into “manual” (skilled/un-skilled) or ”non-manual” (professional/managerial/ intermediate/non-manual skilled) according to UK General Register Office occupational codes (Categories I to III/N and III/M to V respectively).^26^ Since 2008, the three-class version of the National Statistics Socioeconomic classification (NS-SEC) was used (managerial/administrative/professional; intermediate; or routine/manual).^27^ To create a binary “occupation” variable for the whole dataset, the two higher NS-SEC categories were further combined into “non-routine/non-manual” and both variables subsequently merged.

Education was recorded since 2004 and dichotomized into “lower education” (no formal education, primary, lower secondary education) and “higher education” (Upper secondary, post-secondary non-tertiary, tertiary education).

### Outcomes

Pre-stroke risk factor diagnoses were collected as binary variables from hospital/GP records, including hypertension, hypercholesterolaemia (from 1997), diabetes mellitus, myocardial infarction, atrial fibrillation (AF), and body mass index (BMI≥25 categorized as overweight/obese, from 2001). Smoking (current or ex-smoker/never) was obtained from participant interviews. AF newly diagnosed at stroke was collected since 2002.

Pre-stroke risk factor treatments are based on prescribed treatments, collected from hospital/GP records. They include antihypertensives, antiplatelets, anticoagulants, and cholesterol-lowering and diabetes medication. Treatment rates are expressed as percentages of participants diagnosed with the relevant risk factor, e.g. antihypertensive treatment in those diagnosed with hypertension. Additionally, anticoagulation rates of participants with AF and CHA_2_DS_2_-VASc score ≥2 in men and ≥3 in women (“high-risk AF”) were calculated. The outcome variable “at least one untreated risk factor” refers to participants with untreated hypertension, diabetes, hypercholesterolaemia, AF, myocardial infarction, or TIA.

Stroke subtype was stratified into ischaemic and haemorrhagic (primary intracerebral / subarachnoid). Since 1999 the Trial of Org 10172 in Acute Stroke Treatment (TOAST) classification was collected.^28^

### Statistical Analyses

Participants with first stroke between 1995 and 2024 were stratified into three 10-year cohorts by stroke year (1995-2004, 2005-2014, 2015-2024). Characteristics of the study population were stratified by ethnicity, occupation, and education. Categorical variables were summarized as count (percentage), continuous variables as mean (standard deviation [SD]). Bivariate analyses were performed using chi^2^-test, student’s t-test, and ANOVA as appropriate. P-values for trend over time were calculated using the Cochran-Armitage test for categorical variables and simple linear regression for continuous variables.

Multivariable Poisson regression models with robust standard error were used to estimate adjusted prevalence ratios (aPR) of pre-stroke risk factor diagnoses and treatment rates among ethnic minority versus White participants, those with routine/manual versus non-routine/non-manual occupations, and those with lower versus higher education. aPRs were estimated separately for ethnicity (model 1), occupation (model 2), and education (model 3). Models were run for the whole study population and for each cohort. To investigate whether SES explained ethnic inequalities, a second set of models were mutually adjusted for either ethnicity and occupation (model 4), or ethnicity and education (model 5). Finally, interaction terms were included between ethnicity and occupation (model 6), or ethnicity and education (model 7) to examine whether the effect of ethnicity varied by SES, and vice versa. All models were additionally adjusted for age, sex, and year of stroke, including within cohorts.

SES indicators had significant proportions of missing values (occupation: 23.3%, education: 27.3% from 2004). Baseline characteristics of those with and without missing indicator were compared to assess for potential bias (Table S1/Supplement). When occupation or education served as covariate, missing values were included as separate categories, otherwise complete case analyses were carried out.

Data completeness was over 95% for all outcome variables, apart from hypercholesterolaemia (7.7% missing from 1998/collection start), cholesterol lowering treatment (8.5% missing), smoking (9.9%), and BMI (34.0% from 2001; Table S2/Supplement).

All tests were 2-tailed, and P<0.05 was considered statistically significant. Analyses were performed using Stata 18.0 (StataCorp, Texas/USA).

### Patient and public involvement

The SLSR hosts its own Stroke Research Patient and Family group since 2005 which all participants are eligible to join. The group have input into all stages of the project, including setting study aims and ethical aspects of the research. Study outlines are regularly discussed and outcomes presented, including for this study. Patients are also actively involved in the stakeholder group of the overall KCL stroke research programme and the Steering Committee overseeing the project.^24^

## RESULTS

Between 1995 and 2024, a total of 8,643 participants were registered. 128 participants had no ethnicity, education, and occupation recorded and were excluded, leaving 8,515 participants for analysis (Table 1). 5,255 (62%) participants self-reported as White, 1,303 (16%) as Black Caribbean, and 1,122 (13%) as Black African. 3,964 (61%) reported routine/manual occupations and 1,929 (45%) lower education. Black African participants were younger at first stroke (59.6) than Black Caribbean (68.1) and White participants (71.8, Table 2A) with age disparities declining over time (Table S3A/Supplement). Black Caribbean participants had the highest proportion of routine/manual occupation, while Black African participants had the highest proportion of higher education (Table 2A), even after adjusting for age, sex, and stroke year (S4/Supplement).

**Table 1:**
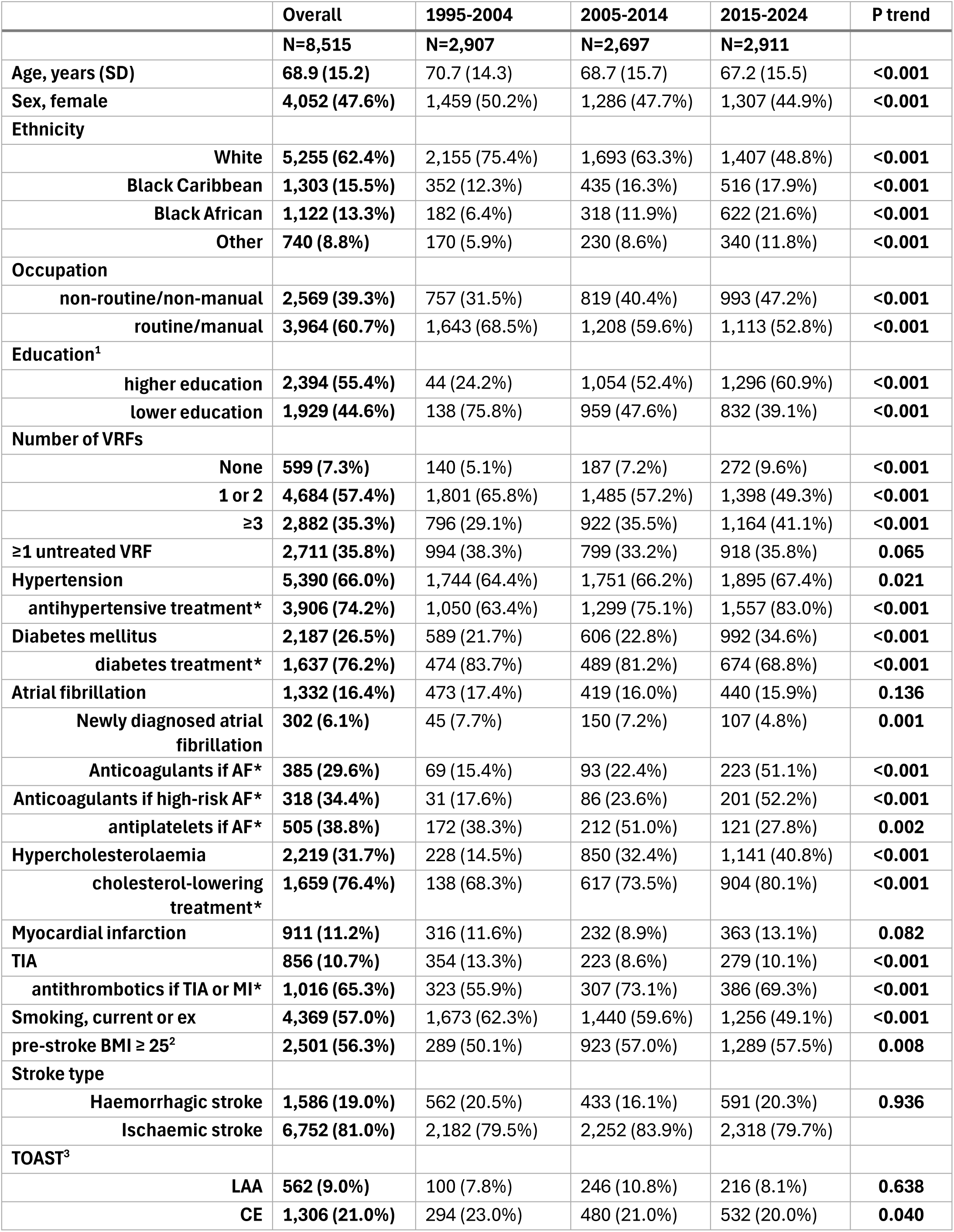
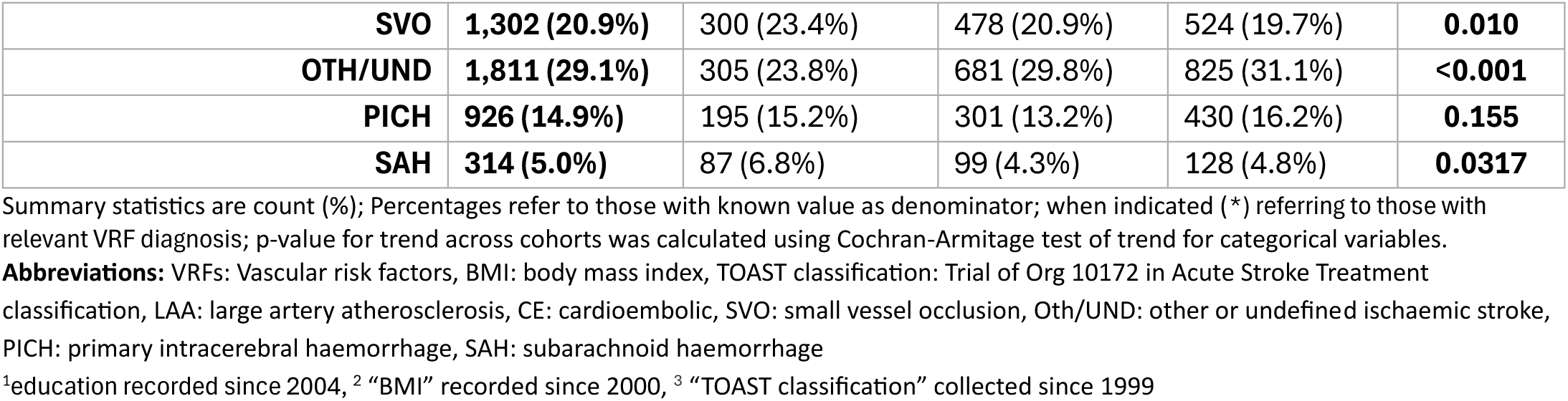
Characteristics of the study population (N=8,515, 1995-2024), overall and by cohorts.

|  | Overall | 1995-2004 | 2005-2014 | 2015-2024 | P trend |
| --- | --- | --- | --- | --- | --- |
|  | **N=8,515** | **N=2,907** | **N=2,697** | **N=2,911** |  |
| **Age, years (SD)** | **68.9 (15.2)** | 70.7 (14.3) | 68.7 (15.7) | 67.2 (15.5) | **<0.001** |
| **Sex, female** | **4,052 (47.6%)** | 1,459 (50.2%) | 1,286 (47.7%) | 1,307 (44.9%) | **<0.001** |
| **Ethnicity** |  |  |  |  |  |
| **White** | **5,255 (62.4%)** | 2,155 (75.4%) | 1,693 (63.3%) | 1,407 (48.8%) | **<0.001** |
| **Black Caribbean** | **1,303 (15.5%)** | 352 (12.3%) | 435 (16.3%) | 516 (17.9%) | **<0.001** |
| **Black African** | **1,122 (13.3%)** | 182 (6.4%) | 318 (11.9%) | 622 (21.6%) | **<0.001** |
| **Other** | **740 (8.8%)** | 170 (5.9%) | 230 (8.6%) | 340 (11.8%) | **<0.001** |
| **Occupation** |  |  |  |  |  |
| **non-routine/non-manual** | **2,569 (39.3%)** | 757 (31.5%) | 819 (40.4%) | 993 (47.2%) | **<0.001** |
| **routine/manual** | **3,964 (60.7%)** | 1,643 (68.5%) | 1,208 (59.6%) | 1,113 (52.8%) | **<0.001** |
| **Education<sup>1</sup>** |  |  |  |  |  |
| **higher education** | **2,394 (55.4%)** | 44 (24.2%) | 1,054 (52.4%) | 1,296 (60.9%) | **<0.001** |
| **lower education** | **1,929 (44.6%)** | 138 (75.8%) | 959 (47.6%) | 832 (39.1%) | **<0.001** |
| **Number of VRFs** |  |  |  |  |  |
| **None** | **599 (7.3%)** | 140 (5.1%) | 187 (7.2%) | 272 (9.6%) | **<0.001** |
| **1 or 2** | **4,684 (57.4%)** | 1,801 (65.8%) | 1,485 (57.2%) | 1,398 (49.3%) | **<0.001** |
| **≥3** | **2,882 (35.3%)** | 796 (29.1%) | 922 (35.5%) | 1,164 (41.1%) | **<0.001** |
| **≥1 untreated VRF** | **2,711 (35.8%)** | 994 (38.3%) | 799 (33.2%) | 918 (35.8%) | **0.065** |
| **Hypertension** | **5,390 (66.0%)** | 1,744 (64.4%) | 1,751 (66.2%) | 1,895 (67.4%) | **0.021** |
| **antihypertensive treatment\*** | **3,906 (74.2%)** | 1,050 (63.4%) | 1,299 (75.1%) | 1,557 (83.0%) | **<0.001** |
| **Diabetes mellitus** | **2,187 (26.5%)** | 589 (21.7%) | 606 (22.8%) | 992 (34.6%) | **<0.001** |
| **diabetes treatment\*** | **1,637 (76.2%)** | 474 (83.7%) | 489 (81.2%) | 674 (68.8%) | **<0.001** |
| **Atrial fibrillation** | **1,332 (16.4%)** | 473 (17.4%) | 419 (16.0%) | 440 (15.9%) | **0.136** |
| **Newly diagnosed atrial fibrillation** | **302 (6.1%)** | 45 (7.7%) | 150 (7.2%) | 107 (4.8%) | **0.001** |
| **Anticoagulants if AF\*** | **385 (29.6%)** | 69 (15.4%) | 93 (22.4%) | 223 (51.1%) | **<0.001** |
| **Anticoagulants if high-risk AF\*** | **318 (34.4%)** | 31 (17.6%) | 86 (23.6%) | 201 (52.2%) | **<0.001** |
| **antiplatelets if AF\*** | **505 (38.8%)** | 172 (38.3%) | 212 (51.0%) | 121 (27.8%) | **0.002** |
| **Hypercholesterolaemia** | **2,219 (31.7%)** | 228 (14.5%) | 850 (32.4%) | 1,141 (40.8%) | **<0.001** |
| **cholesterol-lowering treatment\*** | **1,659 (76.4%)** | 138 (68.3%) | 617 (73.5%) | 904 (80.1%) | **<0.001** |
| **Myocardial infarction** | **911 (11.2%)** | 316 (11.6%) | 232 (8.9%) | 363 (13.1%) | **0.082** |
| **TIA** | **856 (10.7%)** | 354 (13.3%) | 223 (8.6%) | 279 (10.1%) | **<0.001** |
| **antithrombotics if TIA or MI\*** | **1,016 (65.3%)** | 323 (55.9%) | 307 (73.1%) | 386 (69.3%) | **<0.001** |
| **Smoking, current or ex** | **4,369 (57.0%)** | 1,673 (62.3%) | 1,440 (59.6%) | 1,256 (49.1%) | **<0.001** |
| **pre-stroke BMI ≥ 25<sup>2</sup>** | **2,501 (56.3%)** | 289 (50.1%) | 923 (57.0%) | 1,289 (57.5%) | **0.008** |
| **Stroke type** |  |  |  |  |  |
| **Haemorrhagic stroke** | **1,586 (19.0%)** | 562 (20.5%) | 433 (16.1%) | 591 (20.3%) | **0.936** |
| **Ischaemic stroke** | **6,752 (81.0%)** | 2,182 (79.5%) | 2,252 (83.9%) | 2,318 (79.7%) |  |
| **TOAST<sup>3</sup>** |  |  |  |  |  |
| **LAA** | **562 (9.0%)** | 100 (7.8%) | 246 (10.8%) | 216 (8.1%) | **0.638** |
| **CE** | **1,306 (21.0%)** | 294 (23.0%) | 480 (21.0%) | 532 (20.0%) | **0.040** |
| **SVO** | **1,302 (20.9%)** | 300 (23.4%) | 478 (20.9%) | 524 (19.7%) | **0.010** |
| **OTH/UND** | **1,811 (29.1%)** | 305 (23.8%) | 681 (29.8%) | 825 (31.1%) | **<0.001** |
| **PICH** | **926 (14.9%)** | 195 (15.2%) | 301 (13.2%) | 430 (16.2%) | **0.155** |
| **SAH** | **314 (5.0%)** | 87 (6.8%) | 99 (4.3%) | 128 (4.8%) | **0.0317** |

Summary statistics are count (%); Percentages refer to those with known value as denominator; when indicated (\*) referring to those with relevant VRF diagnosis; p-value for trend across cohorts was calculated using Cochran-Armitage test of trend for categorical variables.
**Abbreviations:** VRFs: Vascular risk factors, BMI: body mass index, TOAST classification: Trial of Org 10172 in Acute Stroke Treatment classification, LAA: large artery atherosclerosis, CE: cardioembolic, SVO: small vessel occlusion, Oth/UND: other or undefined ischaemic stroke, PICH: primary intracerebral haemorrhage, SAH: subarachnoid haemorrhage
<sup>1</sup>education recorded since 2004, <sup>2</sup> "BMI" recorded since 2000, <sup>3</sup> "TOAST classification" collected since 1999

**Table 2:**
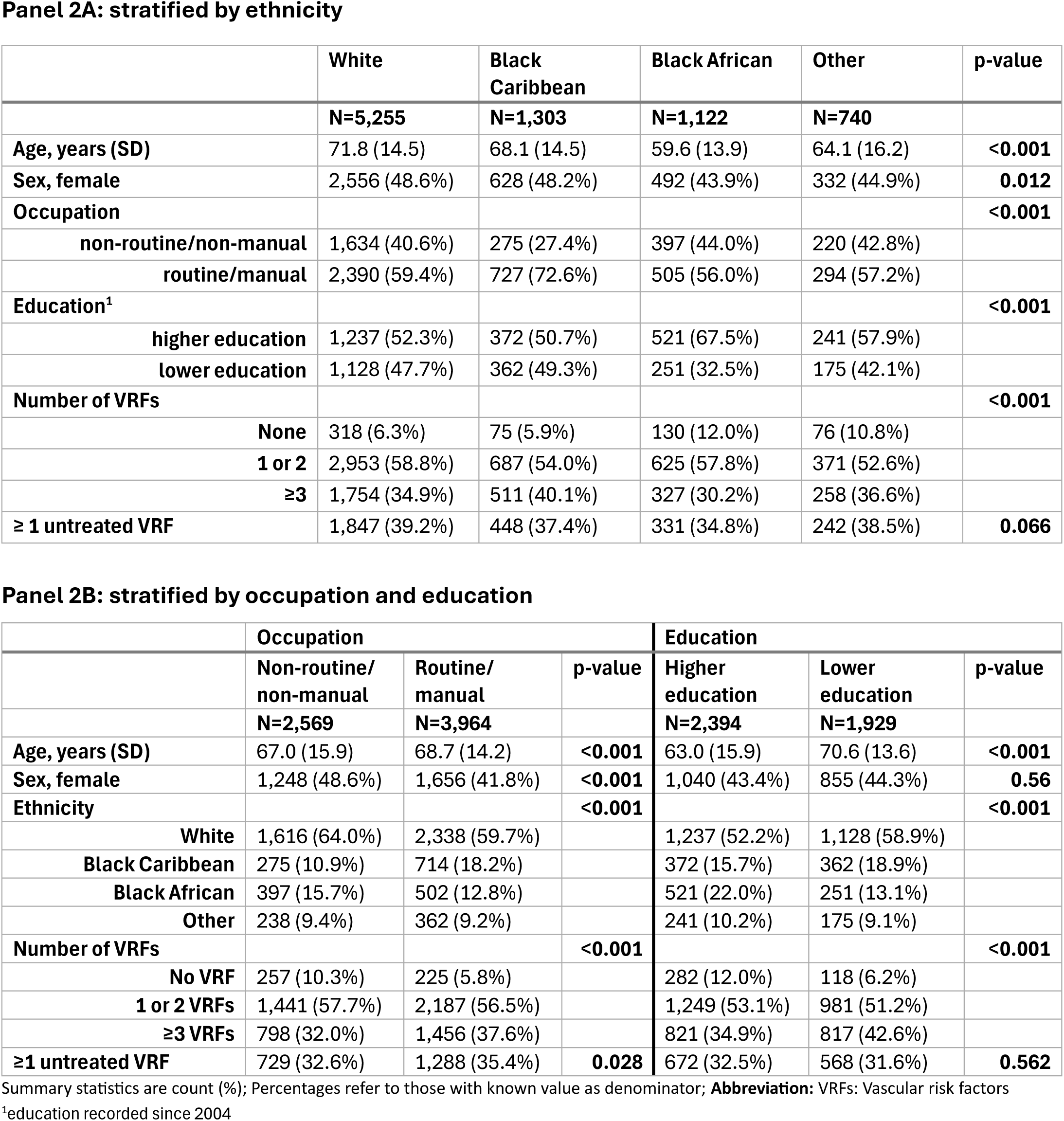
Characteristics of the study population, stratified by sociodemographic categories l 2A: stratified by ethnicity.

The proportion of participants with three or more vascular risk factors increased from 29% in 1995-2004 to 41% in 2015-2024, while the proportion of strokes occurring without pre-stroke risk factor diagnosis also increased from 5% to 10% (Table 1). Prevalence increased most strongly for diabetes (22% to 35%) and hypercholesterolaemia (15% to 41%) but also for hypertension and overweight/obesity, while smoking and TIA declined. The proportion of strokes occurring without prior risk factor diagnosis was higher for Black African participants (12%) than Black Caribbean or White participants (both 6%, Table 2A). Three of more vascular risk factor diagnoses were more common among Black Caribbean participants (40%) than White (35%) or Black African participants (30%). In Black Caribbean and Black African participants this proportion increased more strongly over time (Figure 1). Similarly, lower-SES participants had higher proportions with three of more risk factors than their counterparts (38% vs 32% (occupation) and 43% vs 35% (education), Table 2B), again increasing more strongly over time (Figure 1).

**Figure 1:**
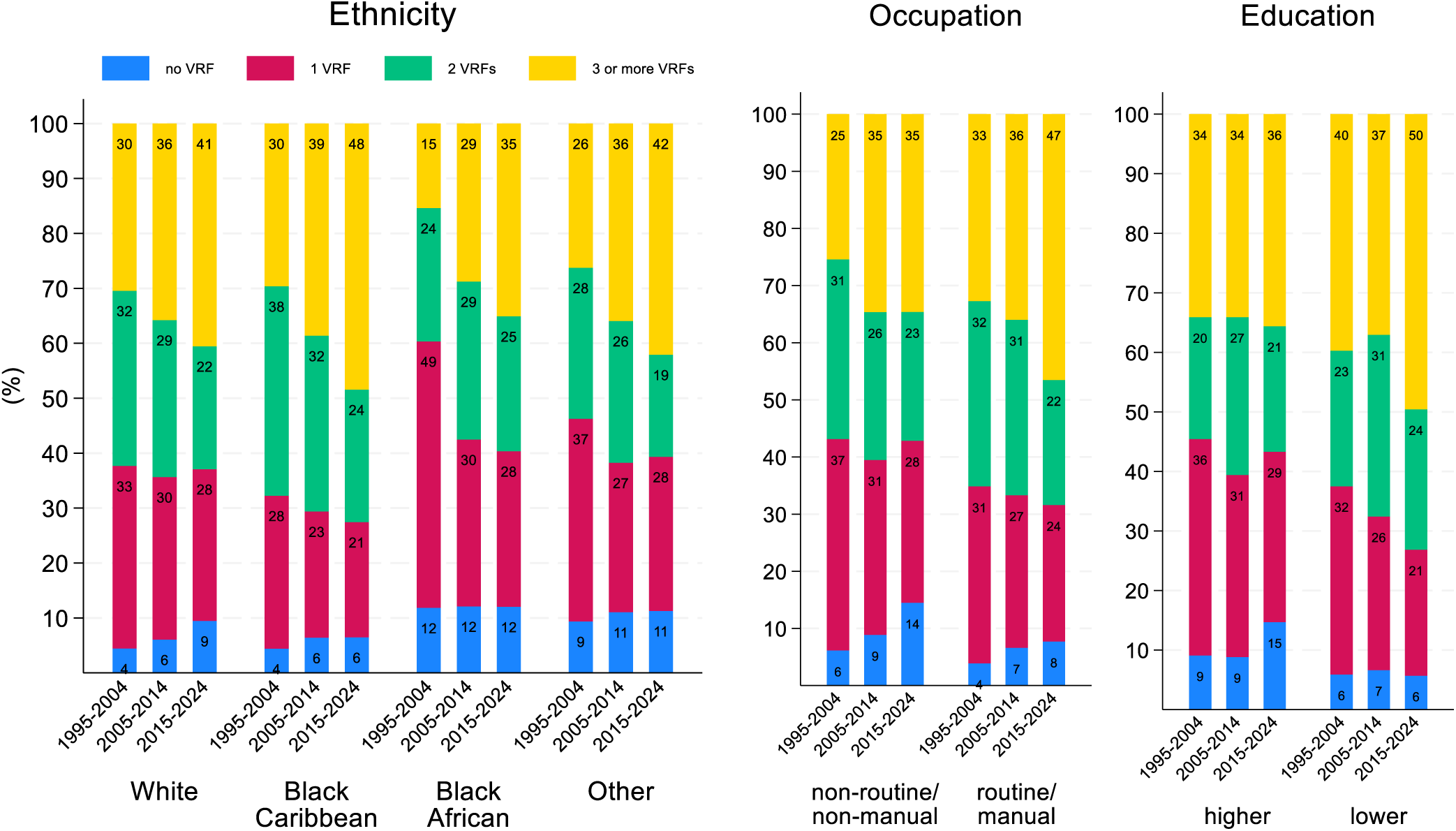
**Trends in the proportion of participants with none, one, two, and three or more pre-stroke vascular risk factors, stratified by ethnicity, occupation, and education, and cohort**

### Risk factor associations and trends

After adjusting for age, sex, and stroke year, vascular risk profiles differed significantly by ethnicity (Figure 2, Model 1). Compared to White participants, diabetes was around twice as common in Black Caribbean (aPR: 2.23 [95%CI:2.04 to 2.43]) and Black African participants (1.92 [1.73 to 2.13]), while hypertension was 29% and 47% and overweight/obesity 13% and 23% more prevalent respectively. Conversely, myocardial infarction, smoking, and AF were markedly less common, with AF prevalence 43% lower in Black Caribbean (aPR 0.57 [0.48 to 0.68]) and 34% lower in Black African participants (0.66 [0.51 to 0.80]).

**Figure 2:**
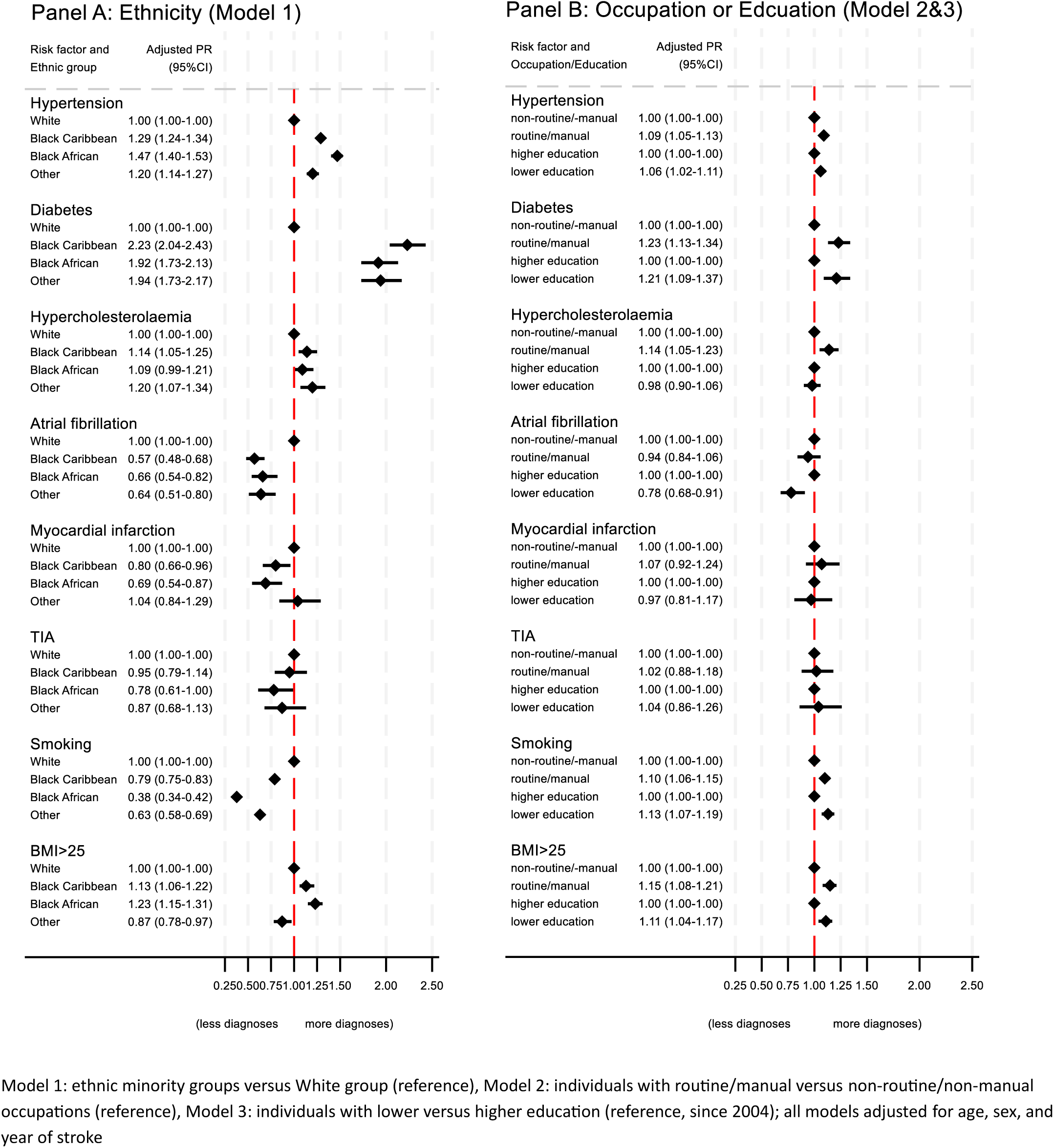
**Adjusted prevalence ratio of pre-stroke vascular risk factor diagnoses between ethnic and socioeconomic groups**

Over time, hypertension declined in White participants, but remained stable in Black Caribbean and increased (non-significantly) in Black African participants (Figure 4). Diabetes increased in all ethnic groups, but this was most pronounced in Black African participants (21% in 1995-2004 to 41% in 2015-2024). In adjusted analyses, associations between Black African ethnicity and hypertension or diabetes became stronger over time (hypertension: aPR 1.37 [1.23 to 1.52] in 1995-2004 versus 1.54 [1.45 to 1.64] in 2015-2024; diabetes: 1.50 [1.09 to 2.07] versus 1.88 [1.65 to 2.15], Table S5/Supplement). AF diagnoses increased in ethnic minority groups (Figure 4) and the strength of the age-adjusted association between lower AF prevalence and Black ethnicity declined but remained significant for Black Caribbean participants (aPR 0.54 [0.29 to 0.73] in 2015-2024).

Diabetes prevalence was 23% higher in participants with routine/manual occupations (vs non-routine/non-manual; aPR 1.23 [1.13 to 1.34]; Figure 2, Model 2) and 21% higher in those with lower education (vs higher education; aPR 1.21 [1.09-1.37]; Model 3). Hypertension, overweight/obesity, and smoking were also significantly more prevalent. Over time, diabetes diagnoses increased in all socioeconomic groups but more strongly in those with routine/manual occupation (23% to 38%) or lower education (20% to 38%, Figure 4). Hypertension diagnoses increased in lower-SES participants but not their counterparts (Figure 4) and associations between hypertension and lower SES became stronger over time (Table S5/Supplement).

Reflecting these risk factor profiles, Black Caribbean, Black African, and lower-SES participants had more strokes due to small vessel occlusion (SVO) than White and higher-SES (Table S6&7/Supplement). Black Caribbean and Black African participants had fewer cardioembolic strokes, but primary intracerebral haemorrhages were most common among Black African participants.

There was only marginal attenuation in risk factor estimates in models mutually adjusted for ethnicity and either occupation or education (Model 4&5, Table S8/Supplement); Further models including interactions between ethnicity and occupation or education were not significant, apart from an interaction between Black African ethnicity and manual/routine occupation being associated with lower hypertension rates and Black Caribbean or African ethnicity and lower education being associated with lower hypertension treatment (Table S9/Supplement).

### Primary prevention – associations and trends

Between 1995-2004 and 2015-2024, treatment rates increased for hypertension (63% to 83%), hypercholesterolaemia (68% to 80%), and AF (anticoagulation: 15 % to 51%), while antidiabetic treatment decreased (84% to 69%, Table 1). Similar trends were observed in all subgroups (Figure 4).

The proportion of stroke patients with at least one diagnosed, untreated pre-stroke vascular risk factor decreased marginally from 38% in 1995-2004 to 36% in 2015-2024 (Table 1). This proportion was higher in White participants (39%) than Black Caribbean (37%) or Black African participants (35%, Table 2A); over time it decreased in White participants from 40% to 35% (p<0.001), increased in Black Caribbean (33% to 40%, p=0.046) and remained stable in Black African participants (33% to 34%, p=0.939, Table S3A/Supplement).

In adjusted analyses, Black Caribbean and Black African participants had higher treatment rates for hypertension and diabetes than White participants (Figure 3), while antithrombotic treatment (in those with previous TIA or MI) was lower in Black African participants (aPR 0.82 [0.70 to 0.95]). All other associations between treatment rates and ethnicity or SES were non-significant. Including ethnicity and education or occupation in mutually adjusted models (Model 4&5), had only marginal impact on the reported associations (Table S8/Supplement).

**Figure 3:**
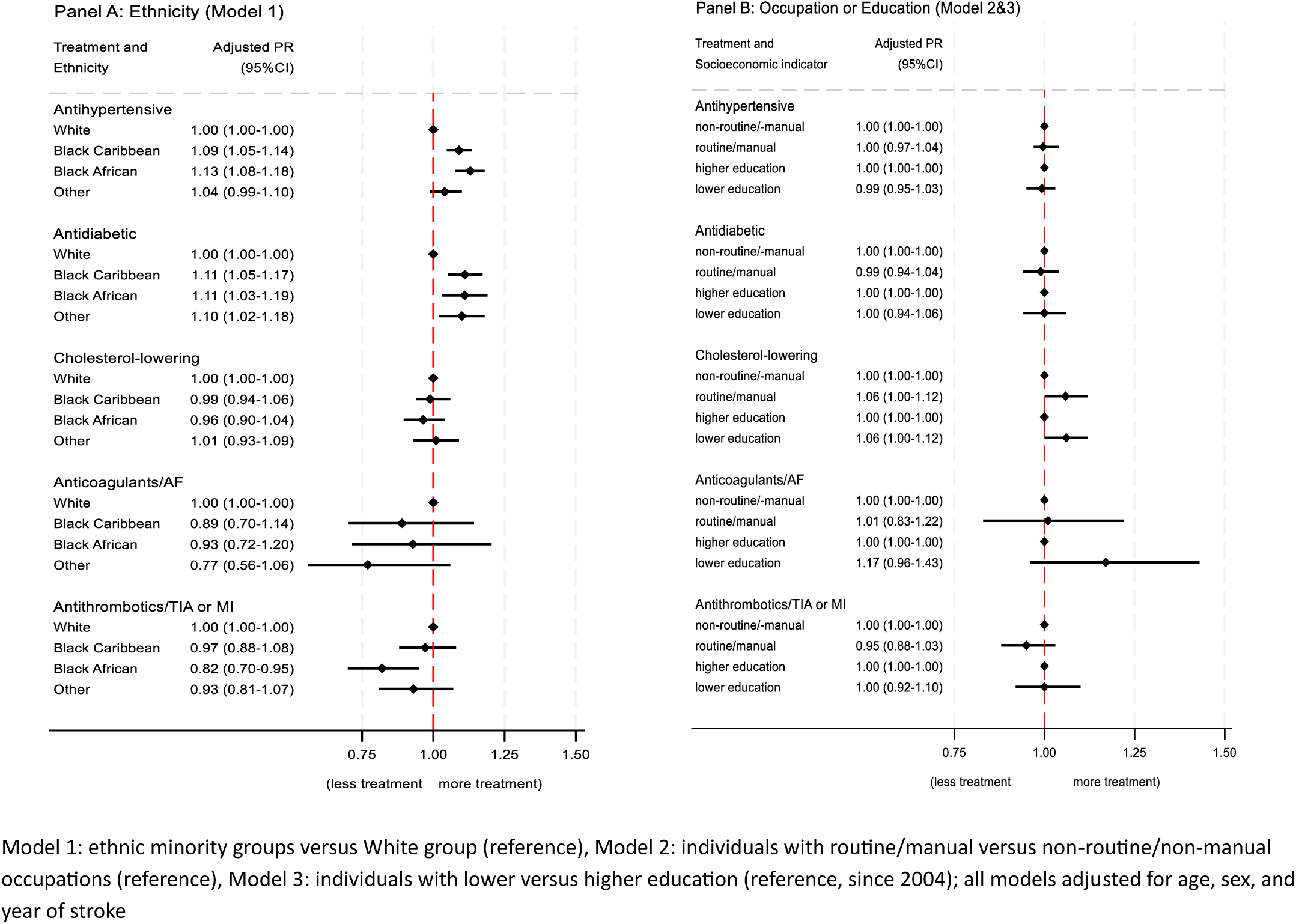
**Adjusted prevalence ratio of pre-stroke treatments in those diagnosed with respective risk factor between ethnic and socioeconomic groups**

**Figure 4:**
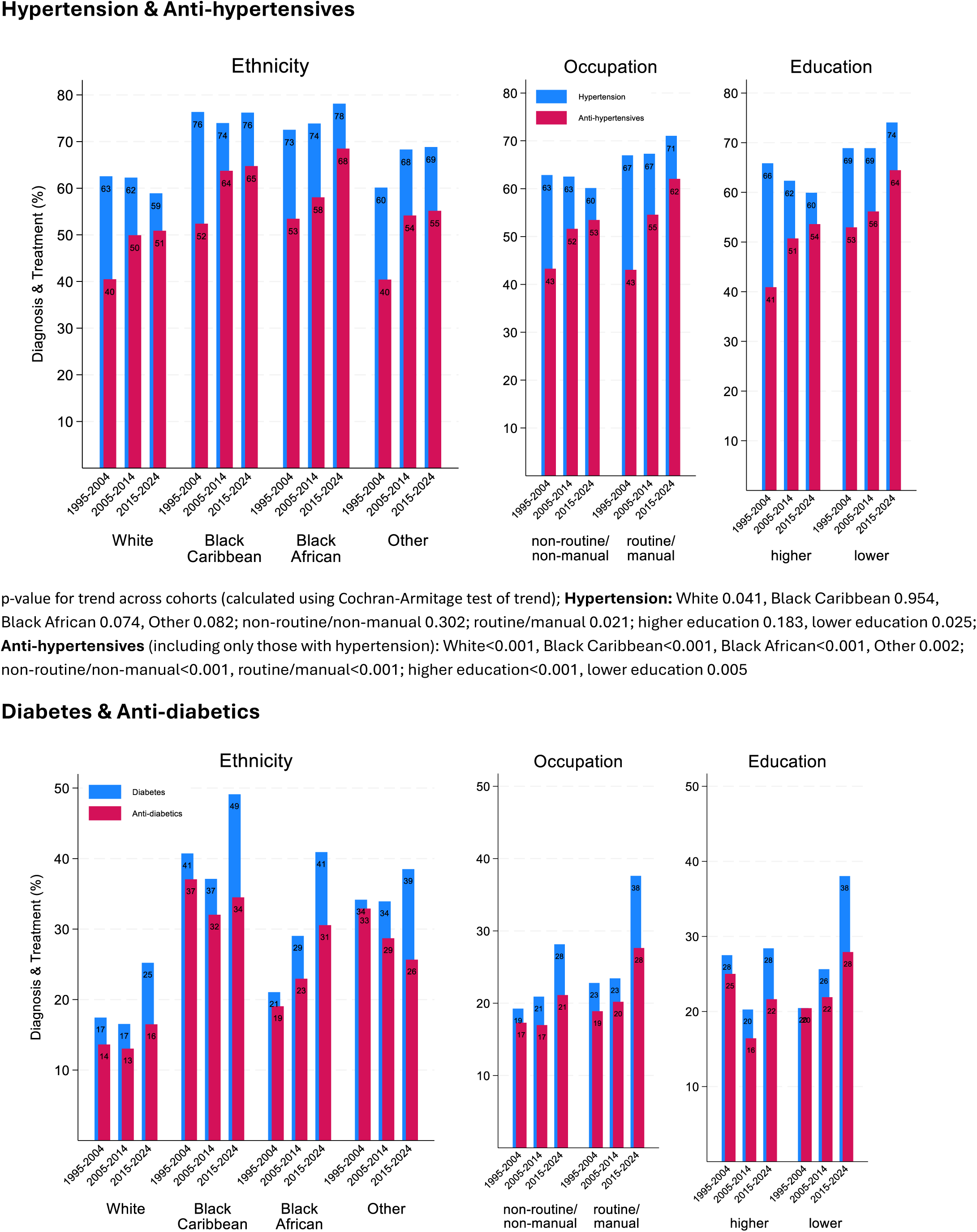
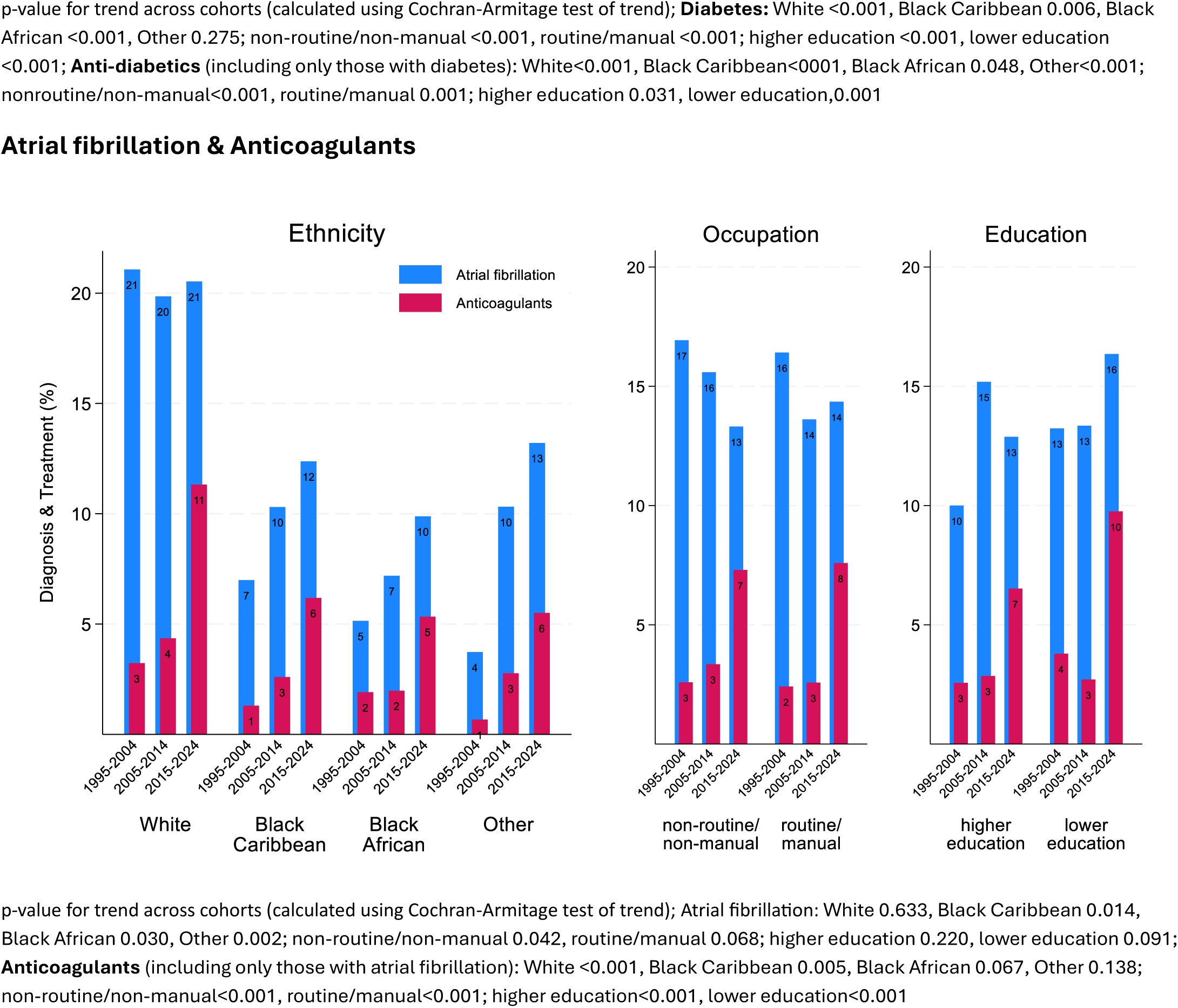
**Trends in pre-stroke vascular risk factor diagnoses and treatment, stratified by ethnicity, occupation, and education**

## DISCUSSION

In this large population-based study of people with stroke spanning three decades, vascular risk factor profiles differed significantly by ethnicity and socioeconomic status; the underlying risk factor burden, particularly hypertension and diabetes, was higher and rising faster in Black Caribbean, Black African, and lower socioeconomic groups, contributing to widening inequalities in stroke incidence. Over a third of strokes continued to occur in people with diagnosed but untreated risk factors and a rising proportion had no pre-stroke risk factor diagnosis, calling for universal as well as targeted improvements in risk factor detection and management.

While ethnic and socioeconomic inequalities in risk factor profiles have been investigated,^3,6,29^ this study adds to the existing evidence in several ways. First, with 30 years of continuous data collection, this study highlights widening inequalities between ethnic and socioeconomic groups, particularly for hypertension and diabetes, thereby identifying targets for primary prevention programmes to reduce health inequalities. Second, while previous studies focused on either ethnic or socioeconomic inequalities, this study includes both and shows that these characteristics, although often similarly associated with risk factors, act independently rather than one driving the other. Third, despite the UK’s universal healthcare provision, pre-stroke risk factor detection and treatment rates in this population-based study remain suboptimal.

In many countries, age-adjusted stroke incidence declined in recent decades due to advances in primary prevention but contrasts with reported increases in vascular risk factor burden.^29–34^ Our study aligns with those reports, showing increasing proportions of participants with three or more risk factors. As well as true increases in prevalence, some increase will be due to improved detection, such as for the observed steep rise in hypercholesterolaemia. Combined with generally improved treatment rates as reported here and incentivised by UK’s pay-for-performance policies,^2^ these might explain declining stroke incidence despite rising risk factor burden. Additionally, improvements in lifestyle factors, e.g. reduced smoking rates, and factors not included here, e.g. improving physical activity levels among UK adults,^30^ might play a role.

However, these overall figures mask large inequalities: we observed higher rates of hypertension and diabetes, prime drivers of stroke incidence inequalities,^6,8,35–37^ in ethnic minority and lower-SES participants and reflected in their higher rates of SVO-related strokes.^38^ These inequalities widened over time, risking further increases in stroke incidence inequalities. Together with higher proportions of strokes without pre-stroke risk factor diagnosis in Black African participants, this demonstrates a need for awareness and screening campaigns targeting high-risk populations. Reducing health inequalities through targeted approaches has been prioritized in both the UK government’s 10-year Health Plan for England^39^ and the national primary care audit CVDPREVENT.^40^ Furthermore, the younger-age onset of hypertension and diabetes^41^ and subsequently first stroke in Black people must be considered in future prevention policies. The current start of vascular health checks in the UK at age 40 might be too late and miss primary prevention opportunities in high-risk groups. Higher prevalence of overweight/obesity in ethnic minority and lower-SES participants additionally underlines the importance of lifestyle and behavioural approaches.

We observed mostly equitable treatment rates between ethnic and socioeconomic groups in line with several^11,42,43^ but not all^44,45^ previous studies. The UK’s universal healthcare provision might play a role, lowering direct financial treatment costs; And the observed higher treatment rates for hypertension and diabetes in Black African and Caribbean participants could reflect higher severity levels of those risk factors. However, while we had no data on risk factor severity or control rates, inequalities in risk factor control have been reported, including specifically for this study’s catchment area in London.^11,42,46–50^ The discrepancy between equitable or higher treatment but diverging control rates calls for patient-centred strategies to improve medication adherence and targeted, more intensive monitoring, mindful of any potential structural racism.^51^ Interventions, based on trust-building, culturally-tailored communications, and shared-decision making, have been shown to be effective.^6,51^ For example, a one-year project in London, delivered through primary care, successfully eradicated a 12%-gap in hypertension control between ethnic minority and White patients.^52^

In this study, socioeconomic inequalities did not explain ethnic inequalities, but ethnicity and SES showed independent effects. Previous evidence of SES-effects on ethnic risk factor inequalities is scarce and conflicting.^20,21^ The UK’s universal healthcare provision might reduce the impact of SES on health and healthcare inequalities, such as for the observed equitable treatment rates. However, there were significant and worsening socioeconomic inequalities in risk factor prevalence, suggesting that factors other than direct financial barriers to healthcare access are important, including upstream social determinants of health, e.g. early childhood factors, education, psychosocial stress, housing quality, or environmental exposure.^53^ Additionally, in this study, the association between ethnic minority and socioeconomic status was nuanced: Black African participants had higher and Black Caribbean participants lower proportions of high education and non-routine/non-manual occupations than White participants. Therefore, apart from non-modifiable biological factors, social and cultural rather than socioeconomic factors might be central in driving ethnic risk factor inequalities and need to be identified in future research.

As in previous studies,^54–56^ Black participants had lower rates of AF. This was unlikely due to selective under-diagnosis, as rates of newly diagnosed AF at stroke were similar across groups. Apart from lower AF prevalence in the study’s Black catchment population, there could be ethnic differences in AF-related stroke risk.^57^ Over time, AF became more prevalent in Black participants, calling for AF awareness campaigns among Black people but also healthcare providers, especially given AF’s silent and often paroxysmal nature.^58^

AF management changed significantly during this study: a 2007 meta-analysis^59^ found anticoagulation superior to aspirin in preventing stroke; DOACs widely replaced Vitamin K antagonists, requiring less monitoring; and the CHA_2_DS_2_-VASc-score superseded the CHADS_2_-score, widening eligibility for anticoagulation. However, despite significant increases, anticoagulation remained widely underused across all groups. While we had no data on contraindications, these are unlikely to justify 48% non-anticoagulated, high-risk people who then proceeded to having stroke. Some previous studies reported lower anticoagulation rates in ethnic minority and lower-SES participants, as well as higher usage of Warfarin compared to DOAC.^60–62^ We found no significant inequalities in anticoagulation rates but could not investigate the relative usage of Warfarin or DOAC due to a lack of data.

### Strengths and Limitations

This study benefits from a continuous 30- year data collection in a population-based cohort facilitating analyses of long-term trends of inequalities in vascular risk factors and treatments, reflecting prevalence trends, diagnostic efforts, and implementation of evidence-based guidelines. Detailed ethnic and socioeconomic data for this large, multi-ethnic cohort enabled us to investigate the effect of SES on ethnic inequalities and a stratification into different Black ethnic groups, often combined in studies and masking distinct cultural identities and migration histories. Additionally, the UK’s universal healthcare provision provides a different lens through which to study health and healthcare inequalities compared to studies based in insurance-based systems.

Limitations include a lack of data on the severity of risk factors, medication adherence, or control rates, which might differ systematically between ethnic and socioeconomic groups and between stroke patients and the general population. Vascular risk factor data included only diagnosed conditions, rather than true prevalence. Additionally, risk factor prevalence in this cohort cannot be generalized to a stroke-free population, despite general trends being broadly similar;^30^ they do however demonstrate important drivers of stroke risk.

Rates of missingness for vascular risk factors and treatments were low but higher for SES indicators. Missingness of SES indicators was not associated with treatment rates but with higher prevalence of pre-stroke hypertension, diabetes, AF, and MI, indicating that we might underestimate their overall prevalence rate.

## Conclusions

Inequalities in risk factor profiles are significant and widening: hypertension and diabetes, major drivers of stroke incidence inequalities, increased disproportionately in ethnic minority and lower-SES participants, calling for targeted primary prevention campaigns. The current start of vascular health checks in the UK at age 40 might miss primary prevention opportunities in high-risk groups with their often-younger age at stroke. One in ten strokes occur without prior risk factor diagnosis, and over a third in people with at least one diagnosed but untreated risk factor, highlighting a need for universal as well as targeted improvements in risk factor detection and management.

## ETHICS STATEMENT

The study has approval from the NHS Health Research Authority (22/WA/0027) and previously from the ethics committees of Guy’s and St Thomas’ Hospital, King’s College Hospital, Queen’s Square, and Westminster Hospital. Informed consent or consultee advice was obtained for all participants.

## TRANSPARENCY STATEMENT

EE affirms that the manuscript is an honest, accurate, and transparent account of the study being reported; that no important aspects of the study have been omitted; and that any discrepancies from the study as originally planned have been explained.

## CONTRIBUTORSHIP STATEMENT

ESE, IJM, and MDLOC conceptualized and designed the study. EE undertook the formal analysis and visualization and wrote the original draft. All other authors reviewed and contributed to subsequent versions. The corresponding author attests that all listed authors meet authorship criteria and that no others meeting the criteria have been omitted.

We thank the stroke survivors, their families and carers, the staff in the stroke services facilitating the study, and all fieldworkers who have collected data over the course of the study.

## DATA AVAILABILITY STATEMENT

Requests for data access for academic use should be made to the SLSR team, where data will be made available subject to academic review and acceptance of a data-sharing agreement.

## Data Availability

All data produced in the present study are available upon reasonable request to the authors.

