## Supplemental Tables for "Ethnic and socioeconomic inequalities in stroke risk factors and primary prevention over three decades: the South London Stroke Register cohort study 1995-2024"

### Supplement

**STROBE Statement—checklist of items that should be included in reports of observational studies . Error! Bookmark not defined.**

**Table S1: Baseline characteristics of the study population with and without missing occupation or education (from 2004)**

| | Occupation | | | Education (from 2004) | | |
|---|---|---|---|---|---|---|
| | Not missing | missing | p-value | Not missing | missing | p-value |
| | N=6,533 | N=1,982 | | N=4,281 | N=1,607 | |
| **Cohort** | | | **<0.001** | | | **<0.001** |
| 1995-2004 | 2,400 (36.7%) | 507 (25.6%) | | 140 (3.3%) | 140 (8.7%) | |
| 2005-2014 | 2,027 (31.0%) | 670 (33.8%) | | 2,013 (47.0%) | 684 (42.6%) | |
| 2015-2024 | 2,106 (32.2%) | 805 (40.6%) | | 2,128 (49.7%) | 783 (48.7%) | |
| **Age, years (SD)** | 68.0 (14.9) | 71.6 (15.9) | **<0.001** | 66.4 (15.4) | 72.3 (15.3) | **<0.001** |
| **Sex, female** | 2,904 (44.5%) | 1,148 (57.9%) | **<0.001** | 1,880 (43.9%) | 847 (52.7%) | **<0.001** |
| **Ethnicity** | | | **<0.001** | | | **<0.001** |
| White | 4,024 (62.5%) | 1,231 (62.2%) | | 2,339 (55.1%) | 950 (59.6%) | |
| Black Caribbean | 1,002 (15.6%) | 301 (15.2%) | | 725 (17.1%) | 267 (16.8%) | |
| Black African | 902 (14.0%) | 220 (11.1%) | | 768 (18.1%) | 195 (12.2%) | |
| Other | 514 (8.0%) | 226 (11.4%) | | 413 (9.7%) | 182 (11.4%) | |
| **Occupation** | | | | | | **0.003** |
| Non-routine/non-manual | 2,569 (39.3%) | | | 1,686 (44.2%) | 205 (37.4%) | |
| Routine/manual | 3,964 (60.7%) | | | 2,132 (55.8%) | 343 (62.6%) | |
| **Education**[1] | | | **<0.001** | | | |
| higher education | 2,176 (56.5%) | 218 (46.5%) | | 2,379 (55.6%) | | |
| lower education | 1,678 (43.5%) | 251 (53.5%) | | 1,902 (44.4%) | | |
| **Number of VRFs** | | | **0.22** | | | **<0.001** |
| no VRF | 482 (7.6%) | 117 (6.5%) | | 399 (9.4%) | 84 (5.7%) | |
| 1 or 2 VRFs | 3,628 (57.0%) | 1,056 (58.6%) | | 2,204 (52.2%) | 830 (56.4%) | |
| more than 2 VRFs | 2,254 (35.4%) | 628 (34.9%) | | 1,623 (38.4%) | 557 (37.9%) | |
| **≥1 untreated VRF** | 2,017 (34.3%) | 694 (41.2%) | **<0.001** | 1,222 (31.9%) | 563 (40.6%) | **<0.001** |
| **Hypertension** | 4,159 (65.5%) | 1,231 (67.6%) | **0.11** | 2,779 (65.5%) | 1,047 (70.0%) | **0.001** |
| antihypertensive treatment* | 3,008 (74.1%) | 898 (74.6%) | **0.71** | 2,195 (79.8%) | 801 (77.6%) | **0.15** |
| **Diabetes mellitus** | 1,638 (25.7%) | 549 (29.3%) | **0.002** | 1,170 (27.6%) | 489 (31.5%) | **0.003** |
| diabetes treatment* | 1,280 (79.6%) | 357 (66.2%) | **<0.001** | 900 (77.9%) | 322 (66.4%) | **<0.001** |
| **Atrial fibrillation** | 952 (15.1%) | 380 (21.1%) | **<0.001** | 598 (14.3%) | 299 (20.3%) | **<0.001** |
| Newly diagnosed atrial fibrillation | 224 (5.9%) | 78 (7.1%) | **0.15** | 180 (5.2%) | 90 (8.0%) | **<0.001** |
| Anticoagulants if AF* | 254 (27.4%) | 131 (34.9%) | **0.007** | 215 (36.3%) | 112 (37.8%) | **0.64** |
| Anticoagulants if high-risk AF* | 206 (32.2%) | 112 (39.2%) | **0.04** | 196 (38.1%) | 100 (37.6%) | **0.9** |
| antiplatelets if AF* | 374 (40.4%) | 131 (34.9%) | **0.067** | 233 (39.3%) | 115 (38.9%) | **0.9** |
| **Hypercholesterolaemia** | 1,684 (31.4%) | 535 (32.7%) | **0.33** | 1,534 (36.4%) | 520 (35.2%) | **0.4** |
| cholesterol-lowering treatment* | 1,254 (76.2%) | 405 (77.0%) | **0.72** | 1,182 (77.9%) | 387 (75.7%) | **0.32** |
| **Myocardial infarction** | 678 (10.7%) | 233 (12.9%) | **0.009** | 425 (10.1%) | 192 (13.1%) | **0.002** |
| **TIA** | 687 (11.0%) | 169 (9.5%) | **0.066** | 390 (9.3%) | 140 (9.6%) | **0.77** |
| antithrombotics in TIA or MI* | 781 (65.1%) | 235 (66.2%) | **0.7** | 527 (72.5%) | 199 (67.0%) | **0.079** |
| **Smoking, current or ex** | 3,672 (58.7%) | 697 (49.8%) | **<0.001** | 2,288 (54.9%) | 560 (53.0%) | **0.27** |
| **pre-stroke BMI≥25**[2] | 2,114 (57.9%) | 387 (49.0%) | **<0.001** | 1,900 (58.9%) | 387 (49.2%) | **<0.001** |
| **Stroke type** | | | **<0.001** | | | **<0.001** |
| Haemorrhagic stroke | 1,101 (17.1%) | 485 (25.3%) | | 658 (15.4%) | 418 (26.2%) | |
| Ischaemic stroke | 5,321 (82.9%) | 1,431 (74.7%) | | 3,617 (84.6%) | 1,177 (73.8%) | |
| **TOAST**[3] | | | **<0.001** | | | **<0.001** |
| LAA | 455 (9.7%) | 107 (7.0%) | | 398 (10.5%) | 87 (6.1%) | |

| | | | | | |
|---|---|---|---|---|---|
| CE | 957 (20.3%) | 349 (23.0%) | | 708 (18.8%) | 359 (25.2%) |
| SVO | 1,104 (23.5%) | 198 (13.0%) | | 881 (23.3%) | 176 (12.4%) |
| OTH/UND | 1,354 (28.8%) | 457 (30.1%) | | 1,172 (31.0%) | 410 (28.8%) |
| PICH | 609 (12.9%) | 317 (20.9%) | | 466 (12.3%) | 302 (21.2%) |
| SAH | 224 (4.8%) | 90 (5.9%) | | 151 (4.0%) | 90 (6.3%) |

Summary statistics are count (%); Percentages refer to those with known value as denominator; when indicated (*) referring to those with relevant VRF diagnosis; p-value for trend across cohorts was calculated using Cochran-Armitage test of trend for categorical variables.
**Abbreviations:** VRFs: Vascular risk factors, BMI: body mass index, TOAST classification: Trial of Org 10172 in Acute Stroke Treatment classification, LAA: large artery atherosclerosis, CE: cardioembolic, SVO: small vessel occlusion, Oth/UND: other or undefined ischaemic stroke, PICH: primary intracerebral haemorrhage, SAH: subarachnoid haemorrhage
[1]education recorded since 2004, [2] “BMI” recorded since 2000, [3] “TOAST classification” collected since 1999

### Table S2: Number of missing values (% of total study population) for each variable

| Variable | N (%) of missing values | collected since |
|---|---|---|
| **Age** | 0 (0.0%) | 1995 |
| **Sex** | 0 (0.0%) | 1995 |
| **Year of stroke** | 0 (0.0%) | 1995 |
| **Ethnicity** | 95 (1.1%) | 1995 |
| **Occupation** | 1,982 (23.3%) | 1995 |
| **Education** | 4,192 (49.2%); 1,607 (27.3%) from 2004 | 2004 |
| **Hypertension** | 348 (4.1%) | 1995 |
| **Antihypertensive treatment** | 335 (4.0%) | 1995 |
| **Diabetes** | 272 (3.2%) | 1995 |
| **Diabetes treatment** | 222 (2.6%) | 1995 |
| **Atrial fibrillation** | 404 (4.7%) | 1995 |
| **Newly diagnosed atrial fibrillation** | 1,499 (23.6%) | 2002 |
| **Anticoagulants** | 318 (3.8%) | 1995 |
| **Antiplatelets** | 318 (3.8%) | 1995 |
| **Hypercholesterolaemia** | 1,522 (17.9%); 7.7% from 1998 | 1998 |
| **Cholesterol-lowering treatment** | 716 (8.5%) | 1995 |
| **Myocardial infarcton** | 401 (4.7%) | 1995 |
| **Transient ischaemic attack** | 388 (4.6%) | 1995 |
| **Smoking** | 836 (9.9%) | 1995 |
| **BMI** | 4,008 (47.6%); 34.0% from 2001 | 2001 |
| **Ischaemic or haemorrhagic stroke** | 177 (2.1%) | 1995 |
| **TOAST classification** | 2,271 (27.0%); 13.7% from 1999 | 1999 |

### Table S3A: Trends in characteristics of the study population, stratified by ethnicity

| | White | | | | Black Caribbean | | | | Black African | | | | Other | | | |
|---|---|---|---|---|---|---|---|---|---|---|---|---|---|---|---|---|
| | **1995-2004** | **2005-2014** | **2015-2024** | **P (trend)** | **1995-2004** | **2005-2014** | **2015-2024** | **P (trend)** | **1995-2004** | **2005-2014** | **2015-2024** | **P (trend)** | **1995-2004** | **2005-2014** | **2015-2024** | **P (trend)** |
| | **N=2,155** | **N=1,693** | **N=1,407** | | **N=352** | **N=435** | **N=516** | | **N=182** | **N=318** | **N=622** | | **N=170** | **N=230** | **N=340** | |
| **Age, years (SD)** | 73.1 (13.3) | 71.6 (15.1) | 69.9 (15.3) | **<0.001** | 66.4 (13.3) | 67.4 (15.0) | 69.8 (14.6) | **<0.001** | 56.4 (14.1) | 58.5 (13.4) | 61.1 (13.9) | **<0.001** | 64.0 (14.4) | 64.9 (16.5) | 63.6 (16.8) | **0.6656** |
| **Sex, female** | 1,118 (51.9%) | 828 (48.9%) | 610 (43.4%) | **<0.001** | 159 (45.2%) | 207 (47.6%) | 262 (50.8%) | **0.100** | 87 (47.8%) | 138 (43.4%) | 267 (42.9%) | **0.299** | 68 (40.0%) | 108 (47.0%) | 156 (45.9%) | **0.277** |
| **Occupation** | | | | | | | | | | | | | | | | |
| higher occupation | 577 (32.8%) | 540 (42.5%) | 517 (52.2%) | **<0.001** | 50 (16.2%) | 91 (27.3%) | 134 (37.1%) | **<0.001** | 70 (47.3%) | 103 (41.9%) | 224 (44.1%) | **0.692** | 42 (31.1%) | 73 (47.1%) | 105 (46.9%) | **0.007** |
| lower occupation | 1,184 (67.2%) | 732 (57.5%) | 474 (47.8%) | **<0.001** | 258 (83.8%) | 242 (72.7%) | 227 (62.9%) | **<0.001** | 78 (52.7%) | 143 (58.1%) | 284 (55.9%) | **0.692** | 93 (68.9%) | 82 (52.9%) | 119 (53.1%) | **0.007** |
| **Education** | | | | | | | | | | | | | | | | |
| higher education | 22 (19.0%) | 613 (49.3%) | 602 (59.8%) | **<0.001** | 10 (33.3%) | 159 (46.9%) | 203 (55.6%) | **0.003** | 8 (44.4%) | 174 (69.0%) | 339 (67.5%) | **0.527** | 4 (23.5%) | 101 (60.1%) | 136 (58.9%) | **0.166** |
| lower education | 94 (81.0%) | 630 (50.7%) | 404 (40.2%) | **<0.001** | 20 (66.7%) | 180 (53.1%) | 162 (44.4%) | **0.003** | 10 (55.6%) | 78 (31.0%) | 163 (32.5%) | **0.527** | 13 (76.5%) | 67 (39.9%) | 95 (41.1%) | **0.166** |
| **≥1 untreated VRF** | 785 (40.4%) | 514 (33.6%) | 432 (35.0%) | **<0.001** | 109 (33.4%) | 125 (31.6%) | 189 (39.6%) | **0.046** | 49 (32.9%) | 97 (36.1%) | 180 (33.7%) | **0.939** | 51 (35.2%) | 63 (32.6%) | 117 (40.2%) | **0.200** |
| **Hypercholesterolaemia** | 156 (14.1%) | 530 (32.2%) | 536 (39.7%) | **<0.001** | 29 (14.6%) | 141 (32.7%) | 231 (46.6%) | **<0.001** | 15 (11.8%) | 97 (31.9%) | 224 (37.4%) | **<0.001** | 24 (23.1%) | 75 (33.5%) | 138 (42.6%) | **<0.001** |
| cholesterol-lowering treatment | 97 (68.8%) | 396 (75.1%) | 431 (80.4%) | **0.002** | 18 (69.2%) | 104 (73.8%) | 186 (80.5%) | **0.069** | 6 (50.0%) | 60 (61.9%) | 179 (79.9%) | **<0.001** | 17 (73.9%) | 57 (76.0%) | 108 (78.3%) | **0.591** |
| **Myocardial infarction** | 264 (13.1%) | 167 (10.2%) | 190 (14.1%) | **0.591** | 22 (6.7%) | 28 (6.7%) | 68 (13.8%) | **<0.001** | 9 (5.2%) | 14 (4.6%) | 53 (8.8%) | **0.190** | 14 (8.7%) | 19 (8.5%) | 49 (15.3%) | **0.015** |
| **TIA** | 287 (14.4%) | 158 (9.6%) | 147 (10.9%) | **<0.001** | 36 (10.8%) | 31 (7.2%) | 61 (12.3%) | **0.329** | 15 (8.6%) | 16 (5.2%) | 44 (7.4%) | **0.952** | 16 (9.9%) | 18 (8.2%) | 27 (8.4%) | **0.617** |
| **Antithrombotics in TIA/ MI** | 277 (57.1%) | 223 (74.3%) | 213 (72.2%) | **<0.001** | 28 (54.9%) | 41 (73.2%) | 78 (70.9%) | **0.075** | 5 (28%) | 20 (67%) | 51 (59%) | **0.072** | 13 (54%) | 23 (68%) | 44 (67%) | **0.349** |
| **Smoking, current or ex** | 1,335 (66.8%) | 1,038 (68.8%) | 777 (63.5%) | **0.115** | 186 (55.9%) | 216 (54.5%) | 218 (48.2%) | **0.028** | 45 (27.4%) | 94 (32.6%) | 128 (22.5%) | **0.029** | 82 (51.9%) | 79 (38.3%) | 119 (41.6%) | **0.074** |
| **pre-stroke BMI≥25** | 188 (46.9%) | 547 (54.6%) | 579 (52.8%) | **0.153** | 52 (57.8%) | 174 (64.7%) | 224 (58.2%) | **0.487** | 34 (72.3%) | 132 (66.7%) | 349 (70.2%) | **0.725** | 15 (40.5%) | 65 (45.1%) | 126 (51.2%) | **0.130** |
| **TOAST** | | | | | | | | | | | | | | | | |
| LAA | 78 (8.8%) | 155 (11.0%) | 132 (10.3%) | **0.329** | 9 (5.6%) | 50 (13.0%) | 27 (5.8%) | **0.224** | 5 (4.3%) | 23 (8.6%) | 39 (6.9%) | **0.688** | 8 (8.9%) | 15 (7.7%) | 18 (5.7%) | **0.225** |
| CE | 233 (26.4%) | 352 (24.9%) | 313 (24.5%) | **0.331** | 27 (16.7%) | 51 (13.2%) | 78 (16.7%) | **0.622** | 13 (11.2%) | 38 (14.1%) | 82 (14.4%) | **0.431** | 8 (8.9%) | 35 (17.9%) | 58 (18.2%) | **0.076** |
| SVO | 178 (20.2%) | 271 (19.2%) | 217 (17.0%) | **0.053** | 62 (38.3%) | 102 (26.4%) | 103 (22.1%) | **<0.001** | 33 (28.4%) | 64 (23.8%) | 133 (23.4%) | **0.327** | 22 (24.4%) | 37 (18.9%) | 60 (18.9%) | **0.332** |
| OTH/UND | 220 (24.9%) | 424 (30.0%) | 373 (29.2%) | **0.052** | 35 (21.6%) | 111 (28.8%) | 163 (34.9%) | **0.001** | 25 (21.6%) | 84 (31.2%) | 180 (31.7%) | **0.072** | 20 (22.2%) | 61 (31.1%) | 99 (31.1%) | **0.78** |
| PICH | 116 (13.1%) | 161 (11.4%) | 187 (14.6%) | **0.204** | 21 (13.0%) | 51 (13.2%) | 74 (15.8%) | **0.262** | 35 (30.2%) | 45 (16.7%) | 106 (18.7%) | **0.045** | 21 (23.3%) | 39 (19.9%) | 62 (19.5%) | **0.483** |
| SAH | 58 (6.6%) | 52 (3.7%) | 57 (4.5%) | **0.045** | 8 (4.9%) | 21 (5.4%) | 22 (4.7%) | **0.794** | 5 (4.3%) | 15 (5.6%) | 28 (4.9%) | **0.961** | 11 (12.2%) | 9 (4.6%) | 21 (6.6%) | **0.225** |

Summary statistics are count (%); Percentages refer to those with known value as denominator; when indicated (*) referring to those with relevant VRF diagnosis; p-value for trend across cohorts was calculated using Cochran-Armitage test of trend for categorical variables.

**Abbreviations:** VRFs: Vascular risk factors, BMI: body mass index, BI: Barthel Index, TOAST classification: Trial of Org 10172 in Acute Stroke Treatment classification, LAA: large artery atherosclerosis, CE: cardioembolic, SVO: small vessel occlusion, Oth/UND: other or undefined ischaemic stroke, PICH: primary intracerebral haemorrhage, SAH: subarachnoid haemorrhage
[1]education recorded since 2004, [2] “BMI” recorded since 2000, [3] “TOAST classification” collected since 1999

### Table S3B: Trends in characteristics of the study population, stratified by occupation and education

| | **Non-routine/non-manual occupation** | | | | **Routine/manual occupation** | | | | **Higher education** | | | | **Lower education** | | | |
|---|---|---|---|---|---|---|---|---|---|---|---|---|---|---|---|---|
| | **1995-2004** | **2005-2014** | **2015-2024** | **P (trend)** | **1995-2004** | **2005-2014** | **2015-2024** | **P (trend)** | **1995-2004** | **2005-2014** | **2015-2024** | **P (trend)** | **1995-2004** | **2005-2014** | **2015-2024** | **P (trend)** |
| | **N=757** | **N=819** | **N=993** | | **N=1,643** | **N=1,208** | **N=1,113** | | N=44 | N=1,054 | N=1,296 | | N=138 | N=959 | N=832 | |
| **Age, years (SD)** | 69.1 (16.1) | 67.5 (16.1) | 65.0 (15.3) | **<0.001** | 71.1 (12.9) | 67.8 (15.1) | 66.4 (14.6) | **<0.001** | 62.4 (17.4) | 64.3 (16.2) | 61.9 (15.4) | **0.001** | 70.1 (12.9) | 70.9 (13.8) | 70.4 (13.5) | **0.688** |
| **Sex, female** | 413 (54.6%) | 389 (47.5%) | 446 (44.9%) | **<0.001** | 749 (45.6%) | 495 (41.0%) | 412 (37.0%) | **<0.001** | 17 (38.6%) | 444 (42.1%) | 579 (44.7%) | **0.155** | 58 (42.0%) | 472 (49.2%) | 325 (39.1%) | **0.002** |
| **Ethnicity** | | | | | | | | | | | | | | | | |
| White | 577 (78.1%) | 540 (66.9%) | 517 (52.8%) | **<0.001** | 1,184 (73.4%) | 732 (61.1%) | 474 (42.9%) | **<0.001** | 22 (50.0%) | 613 (58.5%) | 602 (47.0%) | **<0.001** | 94 (68.6%) | 630 (66.0%) | 404 (49.0%) | **<0.001** |
| Black Caribbean | 50 (6.8%) | 91 (11.3%) | 134 (13.7%) | **<0.001** | 258 (16.0%) | 242 (20.2%) | 227 (20.6%) | **<0.001** | 10 (22.7%) | 159 (15.2%) | 203 (15.9%) | **0.922** | 20 (14.6%) | 180 (18.8%) | 162 (19.7%) | **0.244** |
| Black African | 70 (9.5%) | 103 (12.8%) | 224 (22.9%) | **<0.001** | 78 (4.8%) | 143 (11.9%) | 284 (25.7%) | **<0.001** | 8 (18.2%) | 174 (16.6%) | 339 (26.5%) | **<0.001** | 10 (7.3%) | 78 (8.2%) | 163 (19.8%) | **<0.001** |
| Other | 42 (5.7%) | 73 (9.0%) | 105 (10.7%) | **0.064** | 93 (5.8%) | 82 (6.8%) | 119 (10.8%) | **0.735** | 4 (9.1%) | 101 (9.6%) | 136 (10.6%) | **0.419** | 13 (9.5%) | 67 (7.0%) | 95 (11.5%) | **0.012** |
| **Occupation** | | | | | | | | | | | | | | | | |
| Higher occupation | 757 (100.0%) | 819 (100.0%) | 993 (100.0%) | **.** | 1,643 (100.0%) | 1,208 (100.0%) | 1,113 (100.0%) | **.** | 29 (74.4%) | 505 (53.3%) | 743 (62.5%) | **0.001** | 29 (21.6%) | 209 (25.4%) | 183 (25.4%) | **0.524** |
| lower occupation | . | . | . | **.** | . | . | . | **.** | 10 (25.6%) | 443 (46.7%) | 446 (37.5%) | **0.001** | 105 (78.4%) | 614 (74.6%) | 538 (74.6%) | **0.524** |
| **Education** | | | | | | | | | | | | | | | | |
| Higher education | 29 (50.0%) | 505 (70.7%) | 743 (80.2%) | **<0.001** | 10 (8.7%) | 443 (41.9%) | 446 (45.3%) | **<0.001** | . | . | . | **.** | . | . | . | **.** |
| Lower education | 29 (50.0%) | 209 (29.3%) | 183 (19.8%) | **<0001** | 105 (91.3%) | 614 (58.1%) | 538 (54.7%) | **<0.001** | 44 (100.0%) | 1,054 (100.0%) | 1,296 (100.0%) | **.** | 138 (100.0%) | 959 (100.0%) | 832 (100.0%) | **.** |
| **≥1 untreated VRF** | 224 (33.3%) | 246 (33.7%) | 259 (30.9%) | **0.294** | 610 (40.4%) | 337 (30.2%) | 341 (33.6%) | **<0.001** | 13 (32.5%) | 312 (33.1%) | 347 (31.9%) | **0.587** | 36 (28.1%) | 277 (31.1%) | 255 (32.7%) | **0.280** |
| **Hypercholesterolaemia** | 56 (12.6%) | 268 (33.4%) | 338 (34.7%) | **<0.001** | 151 (17.6%) | 374 (31.4%) | 497 (45.8%) | **<0.001** | 10 (25.6%) | 355 (34.3%) | 463 (36.2%) | **0.176** | 34 (24.8%) | 295 (31.2%) | 384 (47.2%) | **<0.001** |
| cholesterol-lowering treatment | 33 (68.8%) | 183 (69.6%) | 260 (78.1%) | **0.020** | 93 (67.4%) | 286 (77.1%) | 399 (81.1%) | **0.001** | 9 (90.0%) | 258 (73.3%) | 350 (76.6%) | **0.509** | 26 (78.8%) | 222 (75.5%) | 321 (84.7%) | **0.009** |
| **Myocardial infarction** | 77 (10.8%) | 73 (9.1%) | 99 (10.2%) | **0.730** | 188 (12.1%) | 98 (8.3%) | 143 (13.1%) | **0.641** | 4 (9.5%) | 81 (8.0%) | 142 (11.1%) | **0.019** | 9 (6.6%) | 83 (8.8%) | 111 (13.6%) | **<0.001** |
| **TIA** | 78 (11.2%) | 79 (10.0%) | 98 (10.2%) | **0.549** | 230 (15.0%) | 97 (8.1%) | 105 (9.7%) | **<0.001** | 1 (2.5%) | 71 (6.9%) | 128 (10.2%) | **0.002** | 24 (17.8%) | 90 (9.5%) | 82 (10.1%) | **0.144** |
| **antithrombotics in TIA or MI** | 90 (64.7%) | 100 (71.4%) | 112 (68.3%) | **0.540** | 189 (52.6%) | 134 (73.6%) | 156 (72.2%) | **<0.001** | 2 (40.0%) | 104 (72.2%) | 163 (70.9%) | **0.776** | 25 (78.1%) | 117 (73.1%) | 122 (73.5%) | **0.734** |
| **Smoking, current or ex** | 414 (58.7%) | 437 (56.0%) | 444 (46.1%) | **<0.001** | 1,045 (66.8%) | 760 (64.8%) | 572 (53.3%) | **<0.001** | 24 (54.5%) | 601 (58.5%) | 572 (45.4%) | **<0.001** | 90 (66.7%) | 586 (62.1%) | 447 (55.6%) | **0.001** |

| | | | | | | | | | | | | | | | | |
|---|---|---|---|---|---|---|---|---|---|---|---|---|---|---|---|---|
| **pre-stroke BMI>=25** | 77 (43.3%) | 304 (54.9%) | 457 (56.1%) | **0.010** | 197 (53.2%) | 468 (58.9%) | 611 (65.1%) | **<0.001** | 14 (45.2%) | 415 (56.6%) | 621 (58.8%) | **0.168** | 121 (89.0%) | 848 (88.6%) | 706 (84.9%) | **0.017** |
| **TOAST** | | | | | | | | | | | | | | | | |
| LAA | 22 (6.1%) | 73 (10.5%) | 77 (8.5%) | **0.453** | 60 (8.6%) | 121 (11.7%) | 102 (10.0%) | **0.465** | 3 (7.1%) | 105 (11.7%) | 106 (9.0%) | **0.109** | 13 (10.6%) | 100 (12.5%) | 73 (9.5%) | **0.166** |
| CE | 85 (23.6%) | 154 (22.2%) | 162 (17.9%) | **0.010** | 167 (24.0%) | 192 (18.6%) | 197 (19.4%) | **0.035** | 8 (19.0%) | 174 (19.5%) | 206 (17.5%) | **0.276** | 21 (17.1%) | 151 (18.8%) | 154 (20.0%) | **0.393** |
| SVO | 74 (20.6%) | 150 (21.6%) | 192 (21.2%) | **0.877** | 204 (29.3%) | 241 (23.4%) | 243 (23.9%) | **0.021** | 8 (19.0%) | 199 (22.3%) | 247 (21.0%) | **0.631** | 38 (30.9%) | 209 (26.1%) | 188 (24.4%) | **0.148** |
| OTH/UND | 88 (24.4%) | 207 (29.8%) | 303 (33.4%) | **0.002** | 157 (22.6%) | 308 (29.8%) | 291 (28.7%) | **0.012** | 15 (35.7%) | 281 (31.4%) | 375 (31.9%) | **0.970** | 37 (30.1%) | 242 (30.2%) | 235 (30.5%) | **0.878** |
| PICH | 56 (15.6%) | 80 (11.5%) | 131 (14.5%) | **0.957** | 77 (11.1%) | 129 (12.5%) | 136 (13.4%) | **0.155** | 5 (11.9%) | 98 (11.0%) | 172 (14.6%) | **0.020** | 9 (7.3%) | 83 (10.3%) | 100 (13.0%) | **0.028** |
| SAH | 35 (9.7%) | 30 (4.3%) | 41 (4.5%) | **0.002** | 31 (4.5%) | 41 (4.0%) | 46 (4.5%) | **0.878** | 3 (7.1%) | 37 (4.1%) | 71 (6.0%) | **0.128** | 5 (4.1%) | 17 (2.1%) | 20 (2.6%) | **0.794** |

Summary statistics are count (%); Percentages refer to those with known value as denominator; when indicated (*) referring to those with relevant VRF diagnosis; p-value for trend across cohorts was calculated using Cochran-Armitage test of trend for categorical variables.

**Abbreviations:** VRFs: Vascular risk factors, BMI: body mass index, BI: Barthel Index, TOAST classification: Trial of Org 10172 in Acute Stroke Treatment classification, LAA: large artery atherosclerosis, CE: cardioembolic, SVO: small vessel occlusion, Oth/UND: other or undefined ischaemic stroke, PICH: primary intracerebral haemorrhage, SAH: subarachnoid haemorrhage

[1]education recorded since 2004, [2] "BMI" recorded since 2000, [3] "TOAST classification" collected since 1999

### Table S4: Prevalence ratios of routine/manual occupations and lower education among ethnic minorities vs white individuals

| | Ethnicity | aPR | 95%CI |
|---|---|---|---|
| **Occupation** (routine/manual vs non-routine/non-manual) | Black Caribbean | 1.28 | (1.22 - 1.33) |
| | Black African | 1.07 | (1.00 - 1.15) |
| | Other | 1.04 | (0.96 - 1.13) |
| **Education** (lower vs higher) | Black Caribbean | 1.10 | (1.01 - 1.19) |
| | Black African | 0.89 | (0.79 - 0.99) |
| | Other | 1.04 | (0.93 - 1.18) |

All Poisson regression models additionally adjusted for age, sex, and year of stroke

**Table S5: Adjusted prevalence rate ratio (95%CI) of pre-stroke VRFs among ethnic minority vs White participants (model 1), routine/manual vs non-routine/non-manual (model 2), and those with lower vs higher educational attainment (model 3), stratified by cohort**

| Outcome | Exposure | | 1995-2004 | 2005-2014 | 2015-2024 |
|---|---|---|---|---|---|
| Hypertension | Ethnicity (model 1, vs White) | Black Caribbean | **1.30 (1.21-1.39)** | **1.26** (1.18-1.34) | **1.30** (1.22-1.39) |
| | | Black African | **1.37 (1.23-1.52)** | **1.42** (1.31-1.54) | **1.54** (1.45-1.64) |
| | | Other | **1.06 (0.93-1.21)** | **1.19** (1.09-1.31) | **1.29** (1.19-1.40) |
| | Occupation (model 2, vs non-routine/non-manual) | Routine/manual occupation | **1.06 (0.99-1.13)** | **1.07** (1.01-1.14) | **1.15** (1.08-1.22) |
| | Education (model 3, vs higher education) | Lower education | . | **1.03** (0.97-1.10) | **1.09** (1.03-1.16) |
| Diabetes | Ethnicity (model 1, vs White) | Black Caribbean | **2.50 (2.12-2.95)** | **2.33** (1.97-2.75) | **1.95** (1.72-2.21) |
| | | Black African | **1.50 (1.09-2.07)** | **1.98** (1.59-2.46) | **1.88** (1.65-2.15) |
| | | Other | **2.24 (1.76-2.85)** | **2.17** (1.75-2.70) | **1.67** (1.42-1.96) |
| | Occupation (model 2, vs non-routine/non-manual) | Routine/manual occupation | **1.18 (0.99-1.41)** | **1.14** (0.96-1.35) | **1.31** (1.16-1.48) |
| | Education (model 3, vs higher education) | Lower education | . | **1.33** (1.12-1.58) | **1.14** (1.00-1.29) |
| Hypercholest-erolaemia | Ethnicity (model 1, vs White) | Black Caribbean | **1.03 (0.71-1.49)** | **1.04** (0.89-1.21) | **1.19** (1.07-1.33) |
| | | Black African | **0.78 (0.46-1.31)** | **1.09** (0.91-1.31) | **1.16** (1.02-1.31) |
| | | Other | **1.48 (1.01-2.18)** | **1.07** (0.88-1.30) | **1.23** (1.07-1.42) |
| | Occupation (model 2, vs non-routine/non-manual) | Routine/manual occupation | **1.39 (1.04-1.85)** | **0.95** (0.84-1.08) | **1.27** (1.14-1.41) |
| | Education (model 3, vs higher education) | Lower education | . | **0.91** (0.80-1.04) | **1.09** (0.98-1.22) |
| Atrial fibrillation | Ethnicity (model 1, vs White) | Black Caribbean | **0.45 (0.30-0.67)** | **0.59** (0.44-0.79) | **0.62** (0.48-0.80) |
| | | Black African | **0.51 (0.27-0.97)** | **0.56** (0.37-0.85) | **0.77** (0.59-1.00) |
| | | Other | **0.26 (0.12-0.58)** | **0.61** (0.42-0.90) | **0.85** (0.64-1.15) |
| | Occupation (model 2, vs non-routine/non-manual) | Routine/manual occupation | **0.97 (0.81-1.17)** | **0.90** (0.73-1.10) | **0.96** (0.78-1.19) |
| | Education (model 3, vs higher education) | Lower education | . | **0.77** (0.62-0.96) | **0.87** (0.70-1.08) |
| Myocardial infarction | Ethnicity (model 1, vs White) | Black Caribbean | **0.57 (0.37-0.88)** | **0.69** (0.47-1.02) | **1.01** (0.78-1.30) |
| | | Black African | **0.57 (0.29-1.12)** | **0.55** (0.32-0.96) | **0.79** (0.59-1.07) |
| | | Other | **0.79 (0.47-1.31)** | **0.93** (0.59-1.46) | **1.27** (0.96-1.70) |
| | Occupation (model 2, vs non-routine/non-manual) | Routine/manual occupation | **1.05 (0.82-1.35)** | **0.88** (0.66-1.17) | **1.22** (0.96-1.55) |
| | Education (model 3, vs higher education) | Lower education | . | **1.05** (0.77-1.42) | **0.96** (0.75-1.22) |

| | | | | | |
|---|---|---|---|---|---|
| **TIA** | **Ethnicity (model 1, vs White)** | **Black Caribbean** | **0.81 (0.58-1.12)** | **0.84** (0.58-1.22) | **1.15** (0.87-1.51) |
| | | **Black African** | **0.74 (0.44-1.24)** | **0.76** (0.46-1.27) | **0.81** (0.58-1.14) |
| | | **Other** | **0.79 (0.49-1.28)** | **1.00** (0.63-1.60) | **0.88** (0.59-1.30) |
| | **Occupation (model 2, vs non-routine/non-manual)** | **Routine/manual occupation** | **1.30 (1.02-1.65)** | **0.81** (0.61-1.07) | **0.91** (0.70-1.18) |
| | **Education (model 3, vs higher education)** | **Lower education** | **.** | **1.12** (0.82-1.54) | **0.79** (0.60-1.04) |
| **Smoking** | **Ethnicity (model 1, vs White)** | **Black Caribbean** | **0.79 (0.71-0.86)** | **0.78** (0.71-0.85) | **0.79** (0.72-0.88) |
| | | **Black African** | **0.37 (0.29-0.48)** | **0.44** (0.37-0.51) | **0.35** (0.30-0.41) |
| | | **Other** | **0.71 (0.61-0.82)** | **0.53** (0.45-0.63) | **0.66** (0.58-0.76) |
| | **Occupation (model 2, vs non-routine/non-manual)** | **Routine/manual occupation** | **1.09 (1.02-1.17)** | **1.12** (1.04-1.20) | **1.11** (1.01-1.21) |
| | **Education (model 3, vs higher education)** | **Lower education** | **.** | **1.08** (1.00-1.16) | **1.18** (1.08-1.29) |
| **BMI ≥ 25** | **Ethnicity (model 1, vs White)** | **Black Caribbean** | **1.19 (0.97-1.46)** | **1.16** (1.05-1.29) | **1.11** (1.00-1.23) |
| | | **Black African** | **1.35 (1.08-1.70)** | **1.16** (1.03-1.30) | **1.25** (1.15-1.36) |
| | | **Other** | **0.79 (0.52-1.21)** | **0.80** (0.67-0.97) | **0.92** (0.81-1.05) |
| | **Occupation (model 2, vs non-routine/non-manual)** | **Routine/manual occupation** | **1.33 (1.09-1.62)** | **1.08** (0.98-1.19) | **1.17** (1.08-1.26) |
| | **Education (model 3, vs higher education)** | **Lower education** | **.** | **1.06** (0.96-1.17) | **1.13** (1.05-1.22) |

All Poisson regression models additionally adjusted for age, sex, and year of stroke

### Table S6: Characteristics of the study population, stratified by ethnicity

| | White | Black Caribbean | Black African | Other | p-value |
|---|---|---|---|---|---|
| | **N=5,255** | **N=1,303** | **N=1,122** | **N=740** | |
| **Cohort** | | | | | **<0.001** |
| 1995-2004 | 2,155 (41.0%) | 352 (27.0%) | 182 (16.2%) | 170 (23.0%) | |
| 2005-2014 | 1,693 (32.2%) | 435 (33.4%) | 318 (28.3%) | 230 (31.1%) | |
| 2015-2024 | 1,407 (26.8%) | 516 (39.6%) | 622 (55.4%) | 340 (45.9%) | |
| **Age, years (SD)** | 71.8 (14.5) | 68.1 (14.5) | 59.6 (13.9) | 64.1 (16.2) | **<0.001** |
| **Sex, female** | 2,556 (48.6%) | 628 (48.2%) | 492 (43.9%) | 332 (44.9%) | **0.012** |
| **Occupation** | | | | | **<0.001** |
| non-routine/non-manual | 1,634 (40.6%) | 275 (27.4%) | 397 (44.0%) | 220 (42.8%) | |
| routine/manual | 2,390 (59.4%) | 727 (72.6%) | 505 (56.0%) | 294 (57.2%) | |
| **Education**[1] | | | | | **<0.001** |
| higher education | 1,237 (52.3%) | 372 (50.7%) | 521 (67.5%) | 241 (57.9%) | |
| lower education | 1,128 (47.7%) | 362 (49.3%) | 251 (32.5%) | 175 (42.1%) | |
| **Number of VRFs** | | | | | **<0.001** |
| None | 318 (6.3%) | 75 (5.9%) | 130 (12.0%) | 76 (10.8%) | |
| 1 or 2 | 2,953 (58.8%) | 687 (54.0%) | 625 (57.8%) | 371 (52.6%) | |
| ≥3 | 1,754 (34.9%) | 511 (40.1%) | 327 (30.2%) | 258 (36.6%) | |
| **≥ 1 untreated VRF*** | 1,847 (39.2%) | 448 (37.4%) | 331 (34.8%) | 242 (38.5%) | **0.066** |
| **Hypertension** | 3,084 (61.5%) | 957 (75.5%) | 828 (76.0%) | 473 (66.7%) | **<0.001** |
| antihypertensive treatment* | 2,137 (70.4%) | 741 (78.7%) | 672 (82.0%) | 356 (76.2%) | **<0.001** |
| **Diabetes mellitus** | 975 (19.3%) | 547 (42.9%) | 378 (34.5%) | 260 (36.1%) | **<0.001** |
| diabetes treatment* | 712 (73.6%) | 434 (79.6%) | 290 (76.7%) | 201 (78.5%) | **0.044** |
| **Atrial fibrillation** | 1,026 (20.5%) | 128 (10.2%) | 90 (8.3%) | 71 (10.1%) | **<0.001** |
| Newly diagnosed atrial fibrillation | 212 (7.9%) | 39 (4.5%) | 24 (2.8%) | 27 (5.2%) | **<0.001** |
| anticoagulants if AF* | 277 (27.3% | 44 (34.9%) | 40 (44.4%) | 24 (33.8%) | **0.002** |
| Anticoagulants if high-risk AF* | 223 (32.1%) | 39 (37.1%) | 34 (51.5%) | 22 (36.7%) | **0.014** |
| antiplatelets if AF* | 406 (40.4%) | 46 (36.5%) | 28 (31.1%) | 25 (35.2%) | **0.31** |
| **Hypercholesterolaemia** | 1,222 (29.8%) | 401 (35.6%) | 336 (32.6%) | 237 (36.3%) | **<0.001** |
| cholesterol-lowering treatment* | 924 (76.7%) | 308 (77.4%) | 245 (73.6%) | 182 (77.1%) | **0.61** |
| **Myocardial infarction** | 621 (12.4%) | 118 (9.5%) | 76 (7.1%) | 82 (11.6%) | **<0.001** |
| **TIA** | 592 (11.9%) | 128 (10.2%) | 75 (6.9%) | 61 (8.7%) | **<0.001** |
| Antithrombotics if TIA or MI* | 713 (66.0%) | 147 (67.7%) | 76 (56.7%) | 80 (64.5%) | **0.16** |
| **Smoking status, current or ex** | 3,150 (66.6%) | 620 (52.5%) | 267 (26.2%) | 280 (43.1%) | **<0.001** |
| **pre-stroke BMI ≥ 25**[2] | 1,314 (52.6%) | 450 (60.5%) | 515 (69.4%) | 206 (48.2%) | **<0.001** |
| **Stroke type** | | | | | **<0.001** |
| Haemorrhagic stroke | 857 (16.8%) | 252 (19.6%) | 264 (23.6%) | 196 (26.6%) | |
| Ischaemic stroke | 4,248 (83.2%) | 1,037 (80.4%) | 853 (76.4%) | 542 (73.4%) | |
| **TOAST**[3] | | | | | **<0.001** |
| LAA | 365 (10.2%) | 86 (8.5%) | 67 (7.0%) | 41 (6.8%) | |
| CE | 898 (25.1%) | 156 (15.4%) | 133 (14.0%) | 101 (16.7%) | |
| SVO | 666 (18.6%) | 267 (26.3%) | 230 (24.1%) | 119 (19.7%) | |
| OTH/UND | 1,017 (28.4%) | 309 (30.4%) | 289 (30.3%) | 180 (29.8%) | |
| PICH | 464 (13.0%) | 146 (14.4%) | 186 (19.5%) | 122 (20.2%) | |
| SAH | 167 (4.7%) | 51 (5.0%) | 48 (5.0%) | 41 (6.8%) | |

Summary statistics are count (%); Percentages refer to those with known value as denominator; when indicated (*) referring to those with relevant VRF diagnosis;

**Abbreviations:** VRFs: Vascular risk factors, BMI: body mass index, TOAST classification: Trial of Org 10172 in Acute Stroke Treatment classification, LAA: large artery atherosclerosis, CE: cardioembolic, SVO: small vessel occlusion, Oth/UND: other or undefined ischaemic stroke, PICH: primary intracerebral haemorrhage, SAH: subarachnoid haemorrhage

[1]education recorded since 2004, [2] "BMI" recorded since 2000, [3] "TOAST classification" collected since 1999

### Table S7: Characteristics of the study population, stratified by occupation & education

| | Non-routine/ non-manual | Routine/ manual | p-value | Higher education | Lower education | p-value |
|---|---|---|---|---|---|---|
| | **N=2,569** | **N=3,964** | | **N=2,394** | **N=1,929** | |
| **Cohort** | | | **<0.001** | | | **<0.001** |
| 1995-2004 | 757 (29.5%) | 1,643 (41.4%) | | 44 (1.8%) | 138 (7.2%) | |
| 2005-2014 | 819 (31.9%) | 1,208 (30.5%) | | 1,054 (44.0%) | 959 (49.7%) | |
| 2015-2024 | 993 (38.7%) | 1,113 (28.1%) | | 1,296 (54.1%) | 832 (43.1%) | |
| **Age, years (SD)** | 67.0 (15.9) | 68.7 (14.2) | **<0.001** | 63.0 (15.9) | 70.6 (13.6) | **<0.001** |
| **Sex, female** | 1,248 (48.6%) | 1,656 (41.8%) | **<0.001** | 1,040 (43.4%) | 855 (44.3%) | **0.56** |
| **Ethnicity** | | | **<0.001** | | | **<0.001** |
| White | 1,616 (64.0%) | 2,338 (59.7%) | | 1,237 (52.2%) | 1,128 (58.9%) | |
| Black Caribbean | 275 (10.9%) | 714 (18.2%) | | 372 (15.7%) | 362 (18.9%) | |
| Black African | 397 (15.7%) | 502 (12.8%) | | 521 (22.0%) | 251 (13.1%) | |
| Other | 238 (9.4%) | 362 (9.2%) | | 241 (10.2%) | 175 (9.1%) | |
| **Occupation** | | | **<0.001** | | | **<0.001** |
| non-routine/non-manual | 2,569(100.0%) | 0 (0.0%) | | 1,277 (58.7%) | 421 (25.1%) | |
| routine/manual | 0 (0.0%) | 3,964(100.0%) | | 899 (41.3%) | 1,257 (74.9%) | |
| **Education**[1] | | | **<0.001** | | | **<0.001** |
| higher education | 1,277 (75.2%) | 899 (41.7%) | | 2,394(100.0%) | 0 (0.0%) | |
| lower education | 421 (24.8%) | 1,257 (58.3%) | | 0 (0.0%) | 1,929(100.0%) | |
| **Number of VRFs** | | | **<0.001** | | | **<0.001** |
| No VRF | 257 (10.3%) | 225 (5.8%) | | 282 (12.0%) | 118 (6.2%) | |
| 1 or 2 VRFs | 1,441 (57.7%) | 2,187 (56.5%) | | 1,249 (53.1%) | 981 (51.2%) | |
| ≥3 VRFs | 798 (32.0%) | 1,456 (37.6%) | | 821 (34.9%) | 817 (42.6%) | |
| **≥1 untreated VRF** | 729 (32.6%) | 1,288 (35.4%) | **0.028** | 672 (32.5%) | 568 (31.6%) | **0.562** |
| **Hypertension** | 1,542 (61.7%) | 2,617 (68.1%) | **<0.001** | 1,450 (61.2%) | 1,356 (71.0%) | **<0.001** |
| antihypertensive treatment* | 1,133 (75.4%) | 1,875 (73.3%) | **0.14** | 1,138 (79.4%) | 1,072 (79.7%) | **0.85** |
| **Diabetes mellitus** | 583 (23.3%) | 1,055 (27.3%) | **<0.001** | 592 (25.0%) | 585 (30.7%) | **<0.001** |
| **diabetes treatment*** | 455 (79.4%) | 825 (79.7%) | **0.88** | 451 (77.5%) | 456 (78.6%) | **0.64** |
| **Atrial fibrillation** | 376 (15.2%) | 576 (15.0%) | **0.89** | 325 (13.9%) | 276 (14.6%) | **0.53** |
| d/c Atrial fibrillation | 94 (5.9%) | 130 (5.9%) | **0.98** | 85 (4.4%) | 96 (6.2%) | **0.023** |
| anticoagulants if AF* | 110 (30.1%) | 144 (25.7%) | **0.14** | 109 (34.0%) | 106 (38.5%) | **0.24** |
| anticoagulants if high-risk AF* | 92 (34.6%) | 114 (30.6%) | **0.28** | 97 (36.5%) | 99 (39.3%) | **0.51** |
| antiplatelets if AF* | 143 (39.2%) | 231 (41.2%) | **0.54** | 122 (38.0%) | 113 (41.1%) | **0.44** |
| **Hypercholesterolaemia** | 662 (29.8%) | 1,022 (32.6%) | **0.031** | 828 (35.2%) | 713 (37.6%) | **0.11** |
| cholesterol-lowering treatment* | 476 (73.9%) | 778 (77.7%) | **0.076** | 617 (75.3%) | 569 (80.6%) | **0.014** |
| **Myocardial infarction** | 249 (10.0%) | 429 (11.2%) | **0.13** | 227 (9.7%) | 203 (10.7%) | **0.29** |
| **TIA** | 255 (10.4%) | 432 (11.4%) | **0.24** | 200 (8.6%) | 196 (10.4%) | **0.049** |
| antithrombotic if TIA or MI* | 302 (68.2%) | 479 (63.3%) | **0.086** | 269 (71.0%) | 264 (73.7%) | **0.4** |
| **Smoking, current or ex** | 1,295 (52.9%) | 2,377 (62.4%) | **<0.001** | 1,197 (51.4%) | 1,123 (59.7%) | **<0.001** |
| **pre-stroke BMI ≥ 25**[2] | 838 (54.2%) | 1,276 (60.7%) | **<0.001** | 1,050 (57.7%) | 865 (60.4%) | **0.11** |
| **Stroke type** | | | | | | |
| Haemorrhagic stroke | 469 (18.5%) | 632 (16.3%) | **0.025** | 411 (17.2%) | 250 (13.0%) | **<0.001** |
| Ischaemic stroke | 2,073 (81.5%) | 3,248 (83.7%) | | 1,979 (82.8%) | 1,675 (87.0%) | |
| **TOAST**[3] | | | **0.003** | | | **<0.001** |

| | | | | | |
|---|---|---|---|---|---|
| LAA | 172 (8.8%) | 283 (10.3%) | | 214 (10.1%) | 186 (11.0%) |
| CE | 401 (20.5%) | 556 (20.3%) | | 388 (18.4%) | 326 (19.2%) |
| SVO | 416 (21.2%) | 688 (25.1%) | | 454 (21.5%) | 435 (25.7%) |
| OTH/UND | 598 (30.5%) | 756 (27.6%) | | 671 (31.8%) | 514 (30.3%) |
| PICH | 267 (13.6%) | 342 (12.5%) | | 275 (13.0%) | 192 (11.3%) |
| SAH | 106 (5.4%) | 118 (4.3%) | | 111 (5.3%) | 42 (2.5%) |

Summary statistics are count (%); Percentages refer to those with known value as denominator; when indicated (*) referring to those with relevant VRF diagnosis; **Abbreviations:** VRFs: Vascular risk factors, BMI: body mass index, TOAST classification: Trial of Org 10172 in Acute Stroke Treatment classification, LAA: large artery atherosclerosis, CE: cardioembolic, SVO: small vessel occlusion, Oth/UND: other or undefined ischaemic stroke, PICH: primary intracerebral haemorrhage, SAH: subarachnoid haemorrhage

[1]education recorded since 2004, [2] “BMI” recorded since 2000, [3] “TOAST classification” collected since 1999

**Table S8: Adjusted prevalence ratio (95%CI) for pre-stroke VRFs and appropriate primary prevention treatment, mutually adjusted for ethnicity and occupation (model 4) or ethnicity and education (model 5)**

| | Model | Black Caribbean | Black African | Other | Routine/manual | Lower educational attainment |
|---|---|---|---|---|---|---|
| **Hypertension** | Model 4 | 1.28 (1.23-1.33) | 1.46 (1.40-1.53) | 1.20 (1.14-1.27) | 1.08 (1.04-1.12) | |
| | Model 5 | 1.28 (1.22-1.33) | 1.49 (1.42-1.56) | 1.24 (1.17-1.32) | | 1.08 (1.04-1.13) |
| **Diabetes** | Model 4 | 2.19 (2.01-2.39) | 1.92 (1.72-2.13) | 1.93 (1.72-2.16) | 1.14 (1.05-1.25) | |
| | Model 5 | 2.09 (1.90-2.31) | 1.96 (1.75-2.19) | 1.87 (1.64-2.12) | | 1.22 (1.11-1.34) |
| **Hypercholesterol-aemia** | Model 4 | 1.13 (1.03-1.23) | 1.09 (0.98-1.20) | 1.20 (1.08-1.34) | 1.14 (1.05-1.23) | |
| | Model 5 | 1.12 (1.03-1.23) | 1.13 (1.02-1.25) | 1.19 (1.06-1.33) | | 0.99 (0.92-1.08) |
| **AF** | Model 4 | 0.57 (0.48-0.68) | 0.67 (0.54-0.82) | 0.63 (0.50-0.78) | 0.98 (0.88-1.11) | |
| | Model 5 | 0.64 (0.53-0.77) | 0.68 (0.54-0.84) | 0.72 (0.57-0.91) | | 0.81 (0.70-0.93) |
| **MI** | Model 4 | 0.79 (0.65-0.95) | 0.69 (0.54-0.88) | 1.03 (0.83-1.27) | 1.11 (0.95-1.29) | |
| | Model 5 | 0.87 (0.71-1.07) | 0.71 (0.55-0.92) | 1.14 (0.90-1.44) | | 0.98 (0.82-1.18) |
| **TIA** | Model 4 | 0.94 (0.79-1.13) | 0.78 (0.61-0.99) | 0.89 (0.69-1.14) | 1.03 (0.89-1.19) | |
| | Model 5 | 0.99 (0.80-1.22) | 0.78 (0.59-1.02) | 0.89 (0.66-1.20) | | 1.03 (0.85-1.24) |
| **Smoking** | Model 4 | 0.77 (0.73-0.82) | 0.38 (0.34-0.42) | 0.63 (0.58-0.69) | 1.13 (1.08-1.18) | |
| | Model 5 | 0.79 (0.74-0.84) | 0.38 (0.34-0.43) | 0.61 (0.55-0.68) | | 1.11 (1.05-1.16) |
| **BMI ≥ 25** | Model 4 | 1.12 (1.04-1.20) | 1.21 (1.14-1.29) | 0.88 (0.79-0.97) | 1.12 (1.06-1.19) | |
| | Model 5 | 1.11 (1.03-1.19) | 1.23 (1.15-1.31) | 0.88 (0.79-0.98) | | 1.12 (1.05-1.18) |
| **Antihypertensives treatment*** | Model 4 | 1.09 (1.05-1.14) | 1.12 (1.08-1.18) | 1.05 (0.99-1.11) | 1.00 (0.96-1.03) | |
| | Model 5 | 1.08 (1.04-1.13) | 1.13 (1.09-1.19) | 1.03 (0.97-1.09) | | 1.00 (0.96-1.03) |
| **Diabetes treatment*** | Model 4 | 1.10 (1.04-1.17) | 1.10 (1.02-1.18) | 1.10 (1.03-1.18) | 0.98 (0.93-1.03) | |
| | Model 5 | 1.07 (1.00-1.14) | 1.10 (1.01-1.19) | 1.06 (0.97-1.15) | | 0.99 (0.93-1.05) |
| **Cholesterol-lowering treatment*** | Model 4 | 0.99 (0.93-1.05) | 0.96 (0.90-1.04) | 1.00 (0.93-1.08) | 1.06 (1.00-1.12) | |
| | Model 5 | 0.99 (0.93-1.05) | 0.97 (0.90-1.04) | 1.00 (0.92-1.08) | | 1.06 (1.01-1.12) |
| **Anticoagulants if AF*** | Model 4 | 0.90 (0.70-1.14) | 0.93 (0.72-1.20) | 0.76 (0.55-1.50) | 1.01 (0.83-1.23) | |
| | Model 5 | 0.90 (0.71-1.15) | 0.91 (0.70-1.18) | 0.75 (0.54-1.03) | | 1.23 (1.01-1.50) |
| **Antithrombotics if TIA or MI*** | Model 4 | 0.98 (0.88-1.03) | 0.81 (0.70-0.95) | 0.94 (0.81-1.08) | 0.95 (0.88-1.03) | |
| | Model 5 | 0.99 (0.89-1.10) | 0.86 (0.74-1.01) | 0.96 (0.83-1.11) | | 0.99 (0.90-1.08) |

All Poisson regression models additionally adjusted for age, sex, and year of stroke; reference group for ethnicity: White ethnic group, reference group for occupation: non-routine/non-manual occupation; reference group for education: higher educational attainment; when indicated (*) referring to those with relevant VRF diagnosis; Abbreviations: AF: atrial fibrillation, MI: Myocardial infarction, TIA: transient ischaemic attack, BMI: body mass index

**Table S9: Adjusted prevalence ratio (95%CI) for pre-stroke VRFs and appropriate primary prevention treatment, including interaction terms between ethnicity and occupation (model 6) or ethnicity and education (model 7)**

| | Interaction: Ethnicity # Occupation | | | Interaction: Ethnicity # Education | | |
|---|---|---|---|---|---|---|
| **Outcome** | **Independent variable** | **aPR (95%CI)** | **p-value** | **Independent variable** | **aPR (95%CI** | **p-value** |
| **Hypertension** | Black Caribbean | 1.31 (1.20-1.43) | 0.00 | Black Caribbean | 1.31 (1.21-1.43) | 0.00 |
| | Black African | 1.61 (1.50-1.72) | 0.00 | Black African | 1.61 (1.50-1.73) | 0.00 |
| | Other | 1.24 (1.12-1.38) | 0.00 | Other | 1.30 (1.17-1.44) | 0.00 |
| | Routine/manual | 1.10 (1.04-1.16) | 0.00 | Lower education | 1.09 (1.02-1.16) | 0.01 |
| | Black Caribbean # routine/manual | 0.97 (0.88-1.08) | 0.60 | Black Caribbean # lower education | 0.93 (0.84-1.04) | 0.25 |
| | Black African # routine/manual | 0.91 (0.83-0.99) | 0.04 | Black African # lower education | 0.97 (0.88-1.07) | 0.55 |
| | Other # routine/manual | 0.98 (0.86-1.12) | 0.77 | Other # lower education | 0.99 (0.86-1.13) | 0.83 |
| Diabetes | Black Caribbean | 2.24 (1.85-2.71) | 0.00 | Black Caribbean | 1.97 (1.64-2.37) | 0.00 |
| | Black African | 2.15 (1.80-2.57) | 0.00 | Black African | 2.11 (1.78-2.51) | 0.00 |
| | Other | 2.14 (1.73-2.65) | 0.00 | Other | 1.95 (1.56-2.44) | 0.00 |
| | Routine/manual | 1.16 (1.01-1.33) | 0.03 | Lower education | 1.14 (0.96-1.36) | 0.12 |
| | Black Caribbean # routine/manual | 1.03 (0.83-1.29) | 0.78 | Black Caribbean # lower education | 1.18 (0.93-1.50) | 0.18 |
| | Black African # routine/manual | 0.90 (0.72-1.13) | 0.37 | Black African # lower education | 1.05 (0.82-1.34) | 0.70 |
| | Other # routine/manual | 0.94 (0.72-1.23) | 0.66 | Other # lower education | 0.97 (0.71-1.33) | 0.86 |
| Hypercholesterol-aemia | Black Caribbean | 1.10 (0.91-1.33) | 0.33 | Black Caribbean | 1.05 (0.90-1.22) | 0.51 |
| | Black African | 1.13 (0.96-1.34) | 0.14 | Black African | 1.17 (1.02-1.34) | 0.03 |
| | Other | 1.05 (0.84-1.31) | 0.67 | Other | 1.03 (0.84-1.25) | 0.78 |
| | Routine/manual | 1.13 (1.02-1.26) | 0.02 | Lower education | 0.94 (0.84-1.05) | 0.28 |
| | Black Caribbean # routine/manual | 0.98 (0.78-1.22) | 0.84 | Black Caribbean # lower education | 1.11 (0.90-1.37) | 0.33 |
| | Black African # routine/manual | 0.93 (0.74-1.15) | 0.49 | Black African # lower education | 1.02 (0.82-1.27) | 0.85 |
| | Other # routine/manual | 1.21 (0.92-1.59) | 0.17 | Other # lower education | 1.21 (0.92-1.59) | 0.17 |
| Atrial fibrillation | Black Caribbean | 0.58 (0.39-0.86) | 0.01 | Black Caribbean | 0.74 (0.55-0.99) | 0.05 |
| | Black African | 0.75 (0.54-1.04) | 0.09 | Black African | 0.69 (0.50-0.96) | 0.03 |
| | Other | 0.42 (0.25-0.72) | 0.00 | Other | 0.61 (0.39-0.96) | 0.03 |
| | Routine/manual | 0.99 (0.87-1.13) | 0.88 | Lower education | 0.79 (0.66-0.94) | 0.01 |
| | Black Caribbean # routine/manual | 1.00 (0.63-1.59) | 0.99 | Black Caribbean # lower education | 0.84 (0.54-1.30) | 0.44 |
| | Black African # routine/manual | 0.69 (0.42-1.12) | 0.13 | Black African # lower education | 1.00 (0.59-1.70) | 0.99 |
| | Other # routine/manual | 1.55 (0.80-3.00) | 0.20 | Other # lower education | 1.53 (0.85-2.77) | 0.16 |
| Myocardial infarction | Black Caribbean | 1.04 (0.70-1.53) | 0.86 | Black Caribbean | 0.83 (0.57-1.20) | 0.33 |
| | Black African | 0.65 (0.43-0.99) | 0.05 | Black African | 0.79 (0.55-1.12) | 0.18 |
| | Other | 0.98 (0.64-1.51) | 0.94 | Other | 1.04 (0.69-1.58) | 0.85 |
| | Routine/manual | 1.15 (0.96-1.37) | 0.13 | Lower education | 0.96 (0.76-1.21) | 0.73 |

| | | | | | | |
|---|---|---|---|---|---|---|
| | Black Caribbean # routine/manual | 0.63 (0.40-1.01) | 0.06 | Black Caribbean # lower education | 1.14 (0.69-1.88) | 0.62 |
| | Black African # routine/manual | 1.07 (0.63-1.82) | 0.81 | Black African # lower education | 0.85 (0.47-1.53) | 0.58 |
| | Other # routine/manual | 0.98 (0.56-1.70) | 0.93 | Other # lower education | 1.04 (0.56-1.91) | 0.92 |
| TIA | Black Caribbean | 1.01 (0.68-1.50) | 0.96 | Black Caribbean | 0.85 (0.58-1.26) | 0.43 |
| | Black African | 0.77 (0.52-1.14) | 0.19 | Black African | 0.87 (0.60-1.25) | 0.44 |
| | Other | 1.05 (0.69-1.59) | 0.83 | Other | 1.08 (0.69-1.68) | 0.74 |
| | Routine/manual | 1.09 (0.92-1.30) | 0.33 | Lower education | 1.12 (0.88-1.42) | 0.37 |
| | Black Caribbean # routine/manual | 0.85 (0.54-1.34) | 0.49 | Black Caribbean # lower education | 1.18 (0.71-1.96) | 0.53 |
| | Black African # routine/manual | 0.93 (0.56-1.56) | 0.79 | Black African # lower education | 0.59 (0.31-1.15) | 0.12 |
| | Other # routine/manual | 0.60 (0.32-1.09) | 0.09 | Other # lower education | 0.56 (0.27-1.18) | 0.13 |
| Smoking | Black Caribbean | 0.80 (0.71-0.92) | 0.00 | Black Caribbean | 0.82 (0.74-0.91) | 0.00 |
| | Black African | 0.40 (0.34-0.47) | 0.00 | Black African | 0.37 (0.32-0.44) | 0.00 |
| | Other | 0.62 (0.53-0.73) | 0.00 | Other | 0.64 (0.55-0.75) | 0.00 |
| | Routine/manual | 1.12 (1.07-1.18) | 0.00 | Lower education | 1.10 (1.04-1.17) | 0.00 |
| | Black Caribbean # routine/manual | 0.99 (0.86-1.15) | 0.91 | Black Caribbean # lower education | 0.96 (0.83-1.11) | 0.62 |
| | Black African # routine/manual | 0.98 (0.79-1.23) | 0.89 | Black African # lower education | 1.07 (0.83-1.38) | 0.59 |
| | Other # routine/manual | 1.10 (0.90-1.34) | 0.34 | Other # lower education | 0.94 (0.75-1.18) | 0.59 |
| BMI>=25 | Black Caribbean | 1.03 (0.89-1.19) | 0.70 | Black Caribbean | 1.08 (0.97-1.22) | 0.17 |
| | Black African | 1.21 (1.09-1.35) | 0.00 | Black African | 1.24 (1.13-1.35) | 0.00 |
| | Other | 0.91 (0.76-1.08) | 0.27 | Other | 0.88 (0.75-1.04) | 0.13 |
| | Routine/manual | 1.14 (1.05-1.23) | 0.00 | Lower education | 1.13 (1.04-1.23) | 0.01 |
| | Black Caribbean # routine/manual | 1.05 (0.88-1.24) | 0.59 | Black Caribbean # lower education | 0.95 (0.80-1.11) | 0.50 |
| | Black African # routine/manual | 0.97 (0.85-1.11) | 0.67 | Black African # lower education | 0.98 (0.85-1.12) | 0.73 |
| | Other # routine/manual | 0.95 (0.75-1.19) | 0.64 | Other # lower education | 1.01 (0.81-1.27) | 0.92 |
| Hypertension treatment | Black Caribbean | 1.11 (1.02-1.20) | 0.01 | Black Caribbean | 1.13 (1.06-1.21) | 0.00 |
| | Black African | 1.11 (1.03-1.19) | 0.01 | Black African | 1.18 (1.10-1.26) | 0.00 |
| | Other | 1.05 (0.96-1.16) | 0.29 | Other | 1.06 (0.97-1.17) | 0.21 |
| | Routine/manual | 1.00 (0.95-1.06) | 0.88 | Lower education | 1.06 (0.10-1.12) | 0.06 |
| | Black Caribbean # routine/manual | 0.98 (0.89-1.08) | 0.73 | Black Caribbean # lower education | 0.88 (0.80-0.97) | 0.01 |
| | Black African # routine/manual | 0.99 (0.91-1.09) | 0.91 | Black African # lower education | 0.89 (0.81-0.97) | 0.01 |
| | Other # routine/manual | 0.95 (0.84-1.09) | 0.50 | Other # lower education | 0.89 (0.77-1.02) | 0.10 |
| Diabetes treatment | Black Caribbean | 1.06 (0.95-1.18) | 0.33 | Black Caribbean | 1.05 (0.93-1.18) | 0.45 |
| | Black African | 1.04 (0.94-1.17) | 0.40 | Black African | 1.08 (0.97-1.21) | 0.17 |
| | Other | 1.03 (0.91-1.17) | 0.65 | Other | 1.07 (0.94-1.22) | 0.33 |
| | Routine/manual | 0.94 (0.87-1.02) | 0.14 | Lower education | 1.00 (0.90-1.11) | 0.97 |
| | Black Caribbean # routine/manual | 1.06 (0.93-1.22) | 0.36 | Black Caribbean # lower education | 1.01 (0.86-1.18) | 0.93 |
| | Black African # routine/manual | 1.06 (0.92-1.22) | 0.41 | Black African # lower education | 0.99 (0.83-1.16) | 0.86 |

| | | | | | | |
|---|---|---|---|---|---|---|
| | Other # routine/manual | 1.15 (0.99-1.34) | 0.07 | Other # lower education | 0.97 (0.80-1.17) | 0.75 |
| Cholesterol lowering treatment | Black Caribbean | 0.98 (0.86-1.13) | 0.81 | Black Caribbean | 0.99 (0.89-1.11) | 0.89 |
| | Black African | 0.94 (0.83-1.07) | 0.36 | Black African | 0.98 (0.89-1.09) | 0.74 |
| | Other | 0.96 (0.81-1.13) | 0.61 | Other | 1.04 (0.91-1.19) | 0.53 |
| | Routine/manual | 1.03 (0.96-1.11) | 0.44 | Lower education | 1.08 (1.01-1.16) | 0.03 |
| | Black Caribbean # routine/manual | 1.05 (0.90-1.24) | 0.51 | Black Caribbean # lower education | 1.02 (0.88-1.17) | 0.82 |
| | Black African # routine/manual | 1.04 (0.88-1.22) | 0.68 | Black African # lower education | 0.88 (0.75-1.04) | 0.14 |
| | Other # routine/manual | 1.13 (0.93-1.37) | 0.23 | Other # lower education | 0.92 (0.76-1.10) | 0.34 |
| Anticoagulants in Atrial fibrillation | Black Caribbean | 1.03 (0.66-1.62) | 0.89 | Black Caribbean | 0.92 (0.62-1.34) | 0.65 |
| | Black African | 0.95 (0.62-1.45) | 0.80 | Black African | 0.97 (0.63-1.48) | 0.89 |
| | Other | 0.75 (0.40-1.43) | 0.39 | Other | 0.50 (0.23-1.09) | 0.08 |
| | Routine/manual | 1.03 (0.82-1.30) | 0.80 | Lower education | 1.17 (0.92-1.48) | 0.20 |
| | Black Caribbean # routine/manual | 0.83 (0.47-1.48) | 0.54 | Black Caribbean # lower education | 1.04 (0.63-1.74) | 0.87 |
| | Black African # routine/manual | 1.19 (0.66-2.14) | 0.55 | Black African # lower education | 0.99 (0.57-1.74) | 0.98 |
| | Other # routine/manual | 0.88 (0.36-2.17) | 0.78 | Other # lower education | 1.36 (0.52-3.58) | 0.53 |
| Antithrombotics in TIA or myocardial infarction | Black Caribbean | 0.92 (0.75-1.13) | 0.44 | Black Caribbean | 0.93 (0.77-1.12) | 0.44 |
| | Black African | 0.75 (0.57-0.99) | 0.04 | Black African | 0.80 (0.64-0.99) | 0.04 |
| | Other | 0.92 (0.72-1.18) | 0.53 | Other | 0.86 (0.67-1.10) | 0.23 |
| | Routine/manual | 0.94 (0.85-1.03) | 0.17 | Lower education | 0.95 (0.86-1.06) | 0.39 |
| | Black Caribbean # routine/manual | 1.12 (0.87-1.42) | 0.38 | Black Caribbean # lower education | 1.12 (0.88-1.42) | 0.37 |
| | Black African # routine/manual | 1.02 (0.71-1.47) | 0.92 | Black African # lower education | 1.21 (0.86-1.70) | 0.27 |
| | Other # routine/manual | 1.00 (0.70-1.41) | 0.99 | Other # lower education | 1.03 (0.70-1.49) | 0.90 |

All Poisson regression models additionally adjusted for age, sex, and year of stroke; Abbreviations: TIA : transient ischaemic attack, BMI: body mass index
